# Quantifying patient- and hospital-level antimicrobial resistance dynamics in *Staphylococcus aureus* from routinely collected data

**DOI:** 10.1101/2023.02.15.23285946

**Authors:** Quentin Leclerc, Alastair Clements, Helen Dunn, James Hatcher, Jodi A Lindsay, Louis Grandjean, Gwenan M Knight

**Affiliations:** Centre for Mathematical Modelling of Infectious Diseases, Department of Infectious Disease Epidemiology, Faculty of Epidemiology & Public Health, London School of Hygiene & Tropical Medicine, UK; Antimicrobial Resistance Centre, London School of Hygiene & Tropical Medicine, UK; Institute for Infection & Immunity, St George’s University of London, UK; Great Ormond Street Hospital, UK; Department of Infection, Immunity & Inflammation, Institute of Child Health, University College London, UK

**Author notes:** Epidemiology and Modelling of Bacterial Escape to Antimicrobials, Institut Pasteur, Paris, France (Quentin Leclerc).

## Abstract

Antimicrobial resistance (AMR) to all antibiotic classes has been found in the pathogen *Staphylococcus aureus*. The reported prevalence of these resistances vary, driven by within-host AMR evolution at the patient level, and between-host transmission at the hospital level. Without dense longitudinal sampling, pragmatic analysis of AMR dynamics at multiple levels using routine surveillance data is essential to inform control measures.

We explored *S. aureus* AMR diversity in 70,000 isolates from a UK paediatric hospital between 2000-2020, using electronic datasets containing multiple routinely collected isolates per patient with phenotypic antibiograms, hospitalisation information, and antibiotic consumption.

At the hospital-level, the proportion of isolates that were meticillin-resistant (MRSA) increased between 2014-2020 from 25 to 50%, before sharply decreasing to 30%, likely due to a change in inpatient demographics. Temporal trends in the proportion of isolates resistant to different antibiotics were often correlated in MRSA, but independent in meticillin-susceptible *S. aureus*. Ciprofloxacin resistance in MRSA decreased from 70% to 40% of tested isolates between 2007-2020, likely linked to a national policy to reduce fluoroquinolone usage in 2007. At the patient level, we identified frequent AMR diversity, with 4% of patients ever positive for *S. aureus* simultaneously carrying, at some point, multiple isolates with different resistances. We detected changes over time in AMR diversity in 3% of patients ever positive for *S. aureus*. These changes equally represented gain and loss of resistance.

Within this routinely collected dataset, we found that 65% of changes in resistance within a patient’s *S. aureus* population could not be explained by antibiotic exposure or between-patient transmission of bacteria, suggesting that within-host evolution via frequent gain and loss of AMR genes may be responsible for these changing AMR profiles. Our study highlights the value of exploring existing routine surveillance data to determine underlying mechanisms of AMR. These insights may substantially improve our understanding of the importance of antibiotic exposure variation, and the success of single *S. aureus* clones.

## Introduction

Antimicrobial resistance (AMR) poses a major public health threat [1]. Resistance to all antibiotic classes has been reported in *Staphylococcus aureus*, a major human coloniser and nosocomial pathogen [2–5]. AMR in *S. aureus* is complex, with substantial spatio-temporal diversity in the prevalence of different resistances [6]. This AMR diversity can be driven by *S. aureus* transmission across the population at the hospital-level [7,8], or diversity may reflect within-host bacterial evolution at the patient level [9–14]. Since these dynamics at different scales influence each other, studying them simultaneously is essential to inform control measures [15,16].

The most clinically important type of *S. aureus* AMR diversity is the split between meticillin-susceptible and -resistant *S. aureus* (MSSA and MRSA). Approximately 6% of UK *S. aureus* bloodstream infections in 2021 were caused by MRSA [4]. While MSSA isolates tend to be resistant to penicillin only, MRSA carry an SCC*mec* cassette which gives them resistance to most beta-lactam antibiotics [17]. SCC*mec* movement is rare in *S. aureus* isolates [18–20], hence detection of MRSA in an individual is generally attributed to acquisition of MRSA from an external source, rather than gain of SCC*mec* by MSSA already present. Risk factors for MRSA infection or carriage differ between geographical regions and individual ethnicity [21], and include previous antibiotic use, admission to an intensive care unit, and prolonged hospitalisation [22]. *S. aureus* is divided into clonal complexes (CCs), with CC22 (EMRSA-16) and CC30 (EMRSA-15) being the most common MRSA in the UK [23]. Since CCs evolve independently and carry different AMR genes [24], monitoring their prevalence is also essential to understand AMR diversity in *S. aureus* [25].

Previous studies found that patient-level *S. aureus* AMR diversity is common, both in terms of dual MSSA-MRSA carriage and variations in the unique combination of antibiotic resistances and susceptibilities displayed by bacteria in a single host [9–13]. For example, 21% of children sampled before surgery were found to carry both MRSA and MSSA at the same time [12], and between 6.6% and 30% of adult hospital patients simultaneously carried multiple *S. aureus* with different AMR profiles [10,11,13]. However, these studies were often limited to 100-1000 individuals sampled at one time point and mostly focused on diversity within only MRSA populations. MRSA isolates generally carry more resistance genes than MSSA, which leads us to expect more unique combinations of resistances in MRSA than MSSA subpopulations [26]. Thus, our understanding of the temporal dynamics of individual-level AMR diversity is limited.

Antibiotic use will drive AMR diversity on multiple scales - we expect to see correlations between temporal trends of antibiotic consumption and total AMR at the hospital level [27]. At the patient level, an undetected minority resistant subpopulation of resistant bacteria may only become detectable following antibiotic exposure [28]. Although most patients colonised with MRSA in hospitals acquire the bacteria before hospitalisation [29–31], nosocomial transmission may explain other changes in diversity. Finally, changes in patient-level diversity may be the result of within-host horizontal gene transfer (HGT). AMR genes can be frequently gained and lost by *S. aureus in vivo*, translating to changes in diversity [32].

Determining how these mechanisms drive AMR burden within a hospital (within-host evolution, antibiotic selection and transmission) would enable optimisation of interventions. However, collecting the needed dense individual patient data, as well as environmental sampling is expensive and time consuming, as well as hard to justify routinely as directly supporting clinical decision making. In this work, we propose to explore whether we can use routinely collected patient samples to triangulate within-host dynamics with those seen at the hospital-level accounting for non-universal sampling and changes in sampling over time to generate estimates of the relative contributions of each mechanism.

In this study, we aimed to simultaneously quantify hospital- and patient-level dynamics of *S. aureus* AMR diversity using pseudonymised data routinely collected between 2000 and 2021 at Great Ormond Street Hospital (GOSH), London, UK. By quantifying these dynamics, we aimed to highlight opportunities for control measures, and emphasise the value and limitations of routinely collected data, important in the context of increasing usage of this type of data. We explored hospital stay and antibiotic consumption data to also quantify how potential mechanisms contribute to driving these dynamics of AMR diversity.

## Methods

All the analyses and data processing were conducted in the *R* software [33].

### Data processing

#### Antibiogram data

GOSH is a tertiary hospital specialising in paediatric care, receiving between 30,000 and 40,000 inpatients per year including private patients from overseas, and with more than 383 beds spread out across 39 wards (including 44 beds across three intensive care units) [34,35]. Our patient population was defined as anyone at GOSH who had a swab from which *S. aureus* was isolated at any point between 01/02/2000 and 30/11/2021, as determined by the GOSH diagnostic microbiology laboratory. Since swabs are routinely taken for admission and pre-surgical surveillance as well as for clinical concern, this dataset includes both colonising and infecting isolates. Each *S. aureus* isolate was assigned a unique identification number. We included all isolates labelled “Staphylococcus aureus” or “Meticillin-Resistant Staphylococcus aureus”. We excluded 41 isolates labelled “Staphylococcus sp.”, as this may have included *Staphylococcus* species other than *S. aureus*. For each isolate, we had sample collection date, source (wound, urine etc.), and antibiogram data listing isolates resistant to an antibiotic (“R”), or susceptible (“S”), according to nationally recommended reporting criteria.

We labelled all isolates resistant to either cefoxitin, oxacillin, or flucloxacillin as “Meticillin-Resistant Staphylococcus aureus” (MRSA) [36]. All other isolates were assumed to be meticillin-susceptible (MSSA), including 10,031 isolates with no susceptibility information recorded for any of these three antibiotics.

#### Strain typing data

A separate dataset contained genetic typing information on a subset of the *S. aureus* isolates. Isolates were sent for typing to the national reference laboratory. The GOSH guidelines are to systematically type the first MRSA isolate detected in patients and, since 2011, any *S. aureus* isolate suspected of encoding Panton-Valentine leukocidin (PVL), as this can lead to more severe infection [37]. Isolates were labelled either with spa type [38], multilocus sequence typing (MLST) allelic profiles [39], sequence type (ST) [39], or clonal complex (CC) [40]. We matched all this information to clonal complexes, using the online databases from https://spa.ridom.de [38] and https://pubmlst.org [41]. However, we could not link this information to the antibiogram data above, due to the pseudonymised nature of the data.

#### Patient data

Linked routinely collected data for both in- and out-patients from whom these *S. aureus* isolates had been sampled included information on ethnicity of each patient (16 unique ethnicities). We regrouped these ethnicities in three categories: “White British” (which only included the “White British” ethnicity), “Other groups” (all other ethnicities recorded), and “None reported” (patients with no recorded ethnicity, labelled “Prefer Not To Say” or blank).

Hospital admission data included the start and end date for each ward occupied by a patient. Successive events where a patient left a ward and was admitted to another ward on the same day were grouped together to define single hospitalisation events. Antibiotic usage data specified the date and time when each antibiotic was received and name of antibiotic. We matched antibiotics to their class (e.g. meticillin was matched to the “penicillin” class) using online information [42] (Supplementary Table 1).

### Statistical analyses

Linear regression was used to test (1) if the number of isolates per patient followed a consistent trend over time (years) and significantly differed depending on the type of isolate (MRSA or MSSA); and (2) if the number of susceptibility tests conducted and resistances detected per isolate at the hospital level followed a consistent trend over time (years) and significantly differed depending on the type of isolate (MRSA or MSSA).

Spearman’s correlation was used to test the relationship between the proportions of isolates resistant to different antibiotics over time. For this analysis, we considered that colonising isolates were only those originating from nose or throat swabs, as these are the sites most likely to correspond to colonisations rather than infections [43]. The association between colonising or infecting isolates and absence of any susceptibility test conducted was estimated using a Chi-square test. The association between antibiotic usage and changes in diversity was estimated using a Chi-square test.

### Patient-level diversity

#### MRSA and MSSA diversity

For each patient, we chronologically ordered the isolates collected, and identified instances where the type of isolate changed from MSSA to MRSA or vice versa. We then considered the recorded date when each isolate was collected. If a patient had both at least one MRSA and one MSSA isolate detected on the same day, we considered that this indicated meticillin resistance diversity in their *S. aureus* population.

We then identified changes in patient *S. aureus* populations from MRSA to MSSA or vice versa, and notably events where an MSSA isolate was initially identified, followed later by an MRSA isolate. We filtered these events, keeping only those where both the dates of the initial MSSA isolate and the follow-up MRSA isolate were within the time interval of a single hospitalisation event. We excluded events where the MRSA isolate was detected within 2 days of the MSSA, a commonly used threshold to distinguish between already present and acquired MRSA [8].

To identify potential nosocomial transmission of MRSA in a patient *P*, we considered all wards visited by *P* in the interval between their last MSSA-only swab and their first MRSA-positive swab, and checked if, in that same interval, a positive MRSA swab had been reported for any other patient present in any of these wards at the same time as *P*.

To identify changes induced by antibiotic selection, we checked if any antibiotics had been administered to the patients during the interval between the MSSA-positive and MRSA-positive swab.

#### Detected phenotypic resistance diversity

We then searched for patient-level diversity in detected phenotypic resistances. For this, we compared MRSA and MSSA isolates separately. We removed 10,029 isolates which did not have any antibiotic susceptibility test result recorded, leaving more than 60,000 isolates. Antibiograms of chronologically subsequent isolates within patients were compared, and we noted a change in phenotypic resistances displayed if the susceptibility entry for at least one antibiotic in their successive antibiograms changed. We did not consider a change in antibiotic susceptibility to have occurred if only one of the isolates had a recorded result for that antibiotic, and the other had not been tested for that antibiotic.

The same filtering methodology as described above was used to identify differences in antibiograms generated on the same day, on different days, and during a single hospitalisation period.

To identify changes within single hospitalisation periods which may have been induced by antibiotic selection, we checked if the patient was exposed to any antibiotic in the interval between two differing antibiograms, and if the patient was exposed to an antibiotic of the same class as that of the change in resistance.

## Results

### Dynamics of hospital-level *S. aureus* AMR diversity

#### Incidence of MSSA and MRSA isolates

Overall, we obtained information on 72,207 unique *S. aureus* isolates (51,020 MSSA, 21,187 MRSA) from 22,206 patients at Great Ormond Street Hospital (GOSH) between 01/02/2000 and 30/11/2021. Of these patients, 18,700 (84%) only ever tested positive for MSSA, 2,429 (11%) only for MRSA, and 1,077 (5%) tested positive for both MSSA and MRSA (although not necessarily at the same time). The isolates came from a range of sources (nose, wound, blood etc.), representing both colonising and infecting isolates (Supplementary Table 2, Supplementary Figure 1).

Although the total number of *S. aureus* isolates remained consistent over time (Figure 1a), we note a progressive increase in the proportion of MRSA isolates, with a peak in January 2018 (50% of *S. aureus* isolates, Figure 1b). The number of *S. aureus* isolates decreased sharply in April 2020, aligned with the first lockdown during the COVID-19 pandemic in the UK (Figure 1a). After this, the total number of *S. aureus* isolates increased back to pre-2020 levels, and the proportion of MRSA isolates stabilised around 30% (Figure 1a-b).

**Figure 1:**
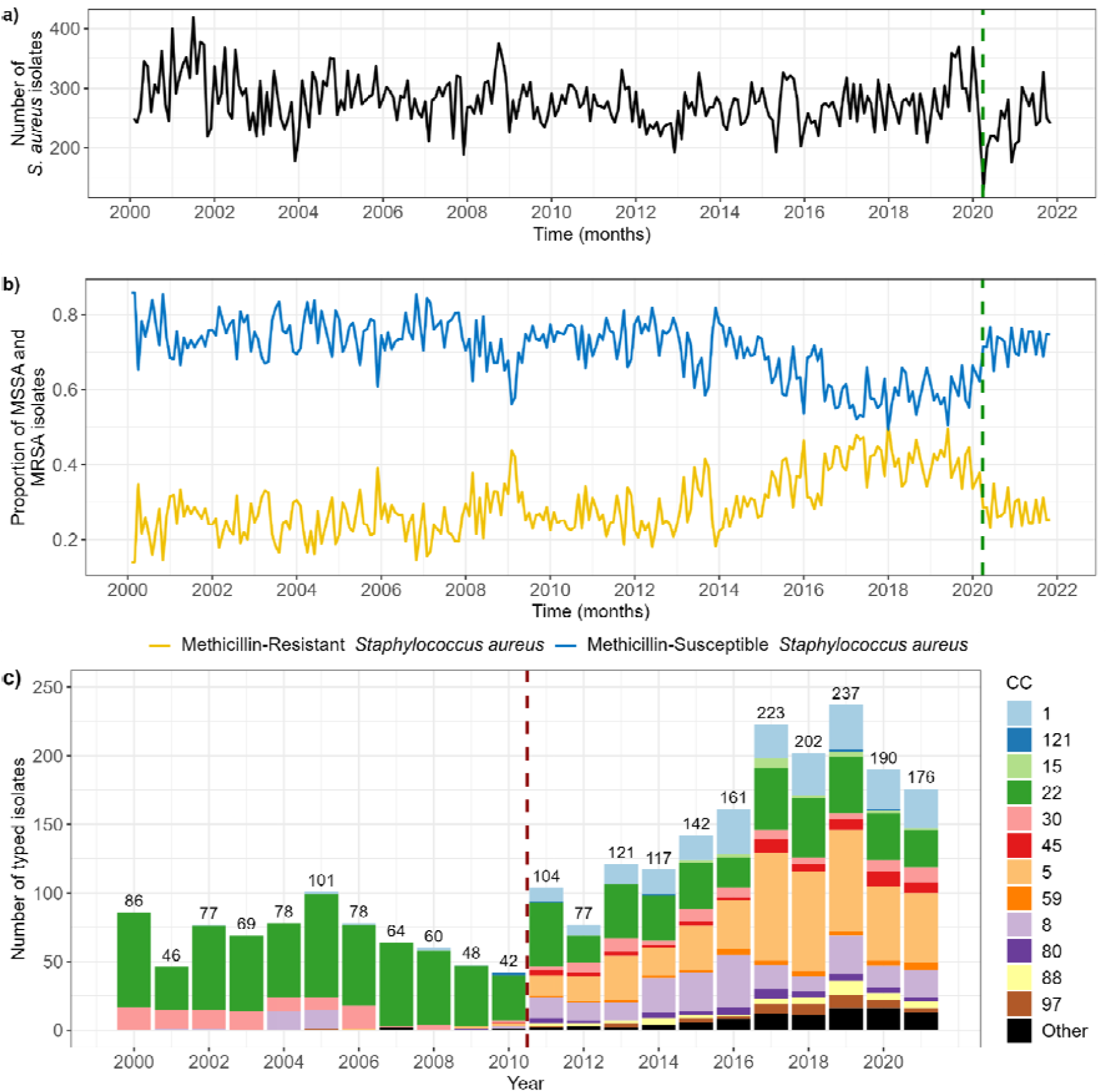
Trends in meticillin-resistant (MRSA) and -susceptible (MSSA) *Staphylococcus aureus* isolates at Great Ormond Street Hospital. **a) Incidence of *S. aureus* isolates. b) Proportion of *S. aureus* isolates which are MRSA or MSSA.** Isolates are grouped by calendar month of date of collection. Vertical green dashed line shows the date when lockdown began in the UK during the first wave of the COVID-19 pandemic (29/03/20). **c) Distribution of *S. aureus* clonal complexes (CC) over time**. Isolates are grouped by calendar year of date of collection. Numbers above each bar denote the total number of typed isolates that year. Only a subset of *S. aureus* patient isolates are typed. Vertical red dashed line shows the year when typing was extended to include isolates suspected of encoding PVL (2011).

These values are not directly equivalent to the number and proportion of unique patients positive for MRSA and/or MSSA, since multiple isolates were frequently recorded per patient (Supplementary Figure 2). More MRSA isolates were always obtained per patient than MSSA (regression coefficient = 1.91, p value < 0.001), and this sampling frequency negligibly varied over time (coeff. = -0.00009, p < 0.001) (Supplementary Figure 2), hence this does not explain variations in the number of MRSA and MSSA isolates seen at the hospital level. However, we found that the increase in proportion of MRSA isolates aligned with an increase in the proportion of patients with an ethnicity other than “White British” admitted to GOSH (Supplementary Figure 3).

The prevalence of different clonal complexes (CCs) changed over time (Figure 1c). Until 2011, CC22 was dominant. After that point, there was a change in guidelines at GOSH to systematically type isolates suspected of encoding PVL, and we observed a substantial increase in the number of CCs reported (up to 12), alongside a progressive increase in CC1 and CC5.

#### Overview of antibiotic susceptibility testing and resistance detection

We obtained antibiogram information for 51 unique antibiotics or combinations of antibiotics (e.g. joint amikacin and flucloxacillin resistance) for our *S. aureus* isolates. Until 2011, more susceptibility tests were conducted for MRSA than for MSSA isolates (Figure 2a, coeff. = 4.04, p < 0.001). After 2010, the number of susceptibility tests for MRSA isolates decreased, with a smaller difference compared to MSSA isolates (Figure 2a, coeff. = 1.78, p < 0.001). The number of antibiotic resistances detected in isolates did not change substantially over time (Figure 2b, coeff. = 0.0002, p < 0.001), but was always higher in MRSA than in MSSA isolates (Figure 2b, coeff. = 2.86, p < 0.001). Resistances in MRSA isolates were often correlated with each other, while resistances in MSSA isolates were generally independent (Supplementary Figures 4 and 5). The most common antibiotic susceptibility tests conducted across the entire time period were similar between MRSA and MSSA isolates (Figure 2c-d), although MRSA were more frequently resistant, consistent with Figure 2b.

**Figure 2:**
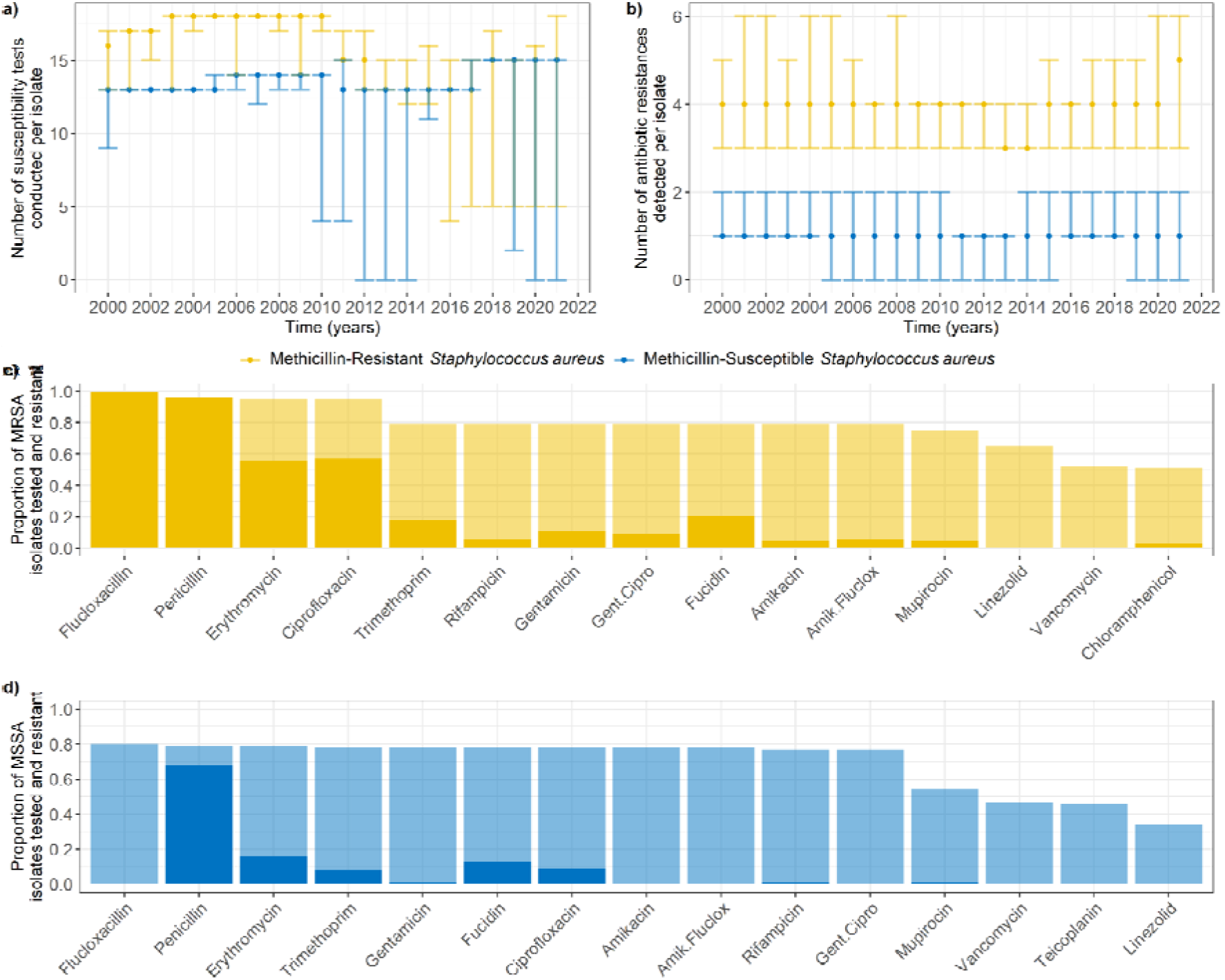
Numbers of susceptibility tests and antibiotic resistances in *S. aureus* isolates. **a) Median number of susceptibility tests conducted per isolate, by year.** Error bars show the interquartile range (first to third quartiles - note these can overlap). **b) Median number of antibiotic resistances detected per isolate, by year**. Error bars show the interquartile range. **c-d) Fifteen most common antibiotic susceptibility tests conducted in meticillin-resistant (c) and -susceptible (d) *S. aureus* isolates, across the entire dataset ordered by number of tests**. Transparent bars indicate the proportion of isolates that were tested for the corresponding antibiotic, and solid bars indicate the proportion of isolates that were resistant. Note that flucloxacillin is an indicator for MRSA, and that we would expect most MSSA and all MRSA isolates to be resistant to penicillin.

Colonising isolates (from nose or throat swabs) were less likely to be tested for antibiotic susceptibility than infecting isolates. Isolates from nose or throat swabs were overrepresented amongst 10,029/72,207 isolates with no recorded susceptibility test (75%), compared to their prevalence amongst all isolates (19%; *X*^2^ = 13,837, df = 1, *p* < 0.001).

#### Trends in antibiotic resistances detected in *S. aureus* isolates

There was substantial variation in resistance trends detected in *S. aureus* (Figure 3, Supplementary Figure 6) as well as substantial variation in the proportions of antibiotics tested for (Figure 4, Supplementary Figure 7).

**Figure 3:**
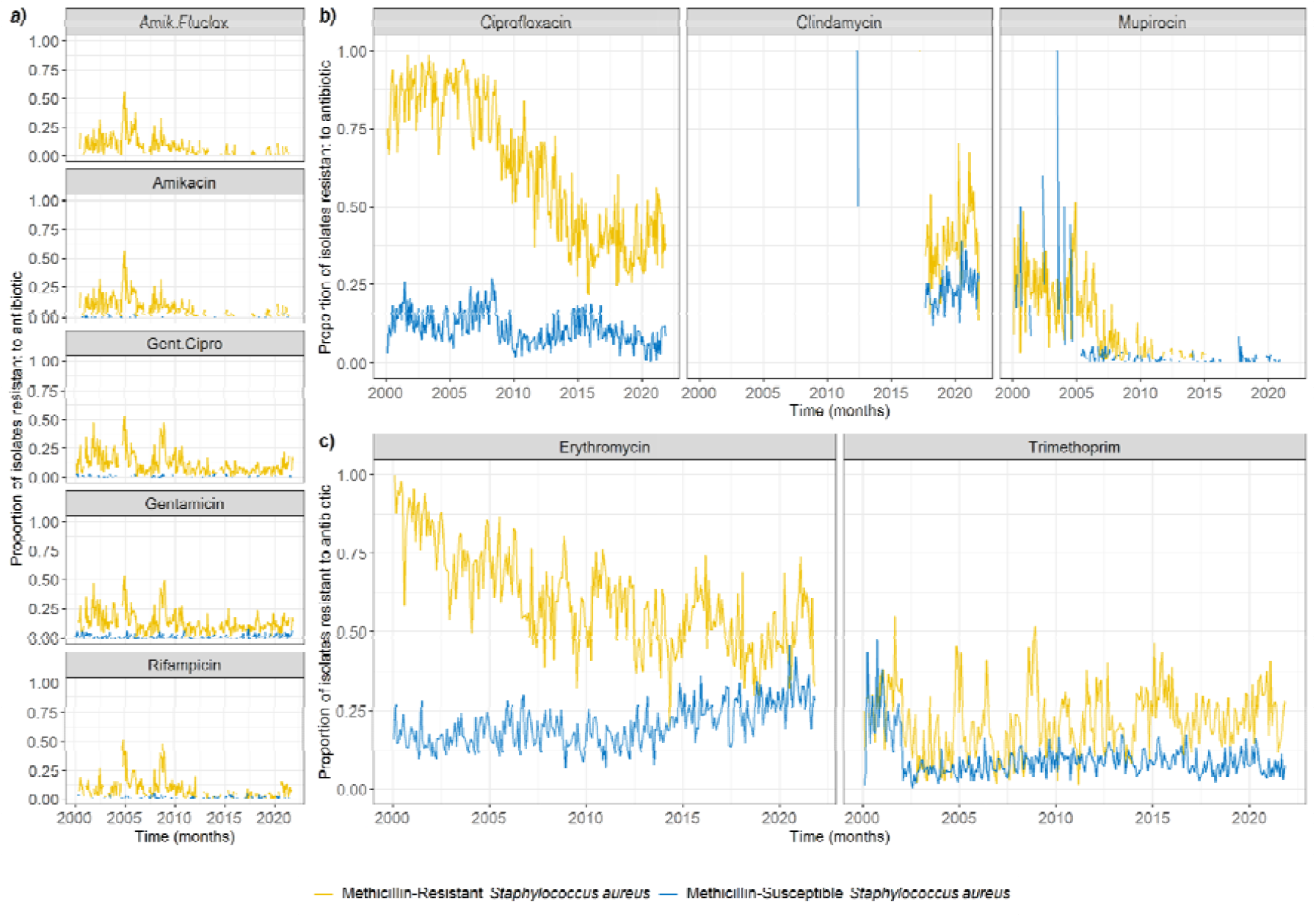
Change in proportion of *S. aureus* isolates resistant to antibiotics over time, out of those tested for resistance to the corresponding antibiotic. These were subjectively grouped in four categories to facilitate visualisation. **a) Strongly correlated antibiotic resistances**. Amik.Fluclox: joint amikacin and flucloxacillin resistance, Gent.Cipro: joint gentamicin and ciprofloxacin resistance. **b) Antibiotic resistances with similar trends between MRSA and MSSA isolates. c) Antibiotic resistances with differing trends between MRSA and MSSA isolates**. Missing lines indicate that no resistance to the corresponding antibiotic was detected at that time (which may be due to no testing having been conducted at that time).

**Figure 4:**
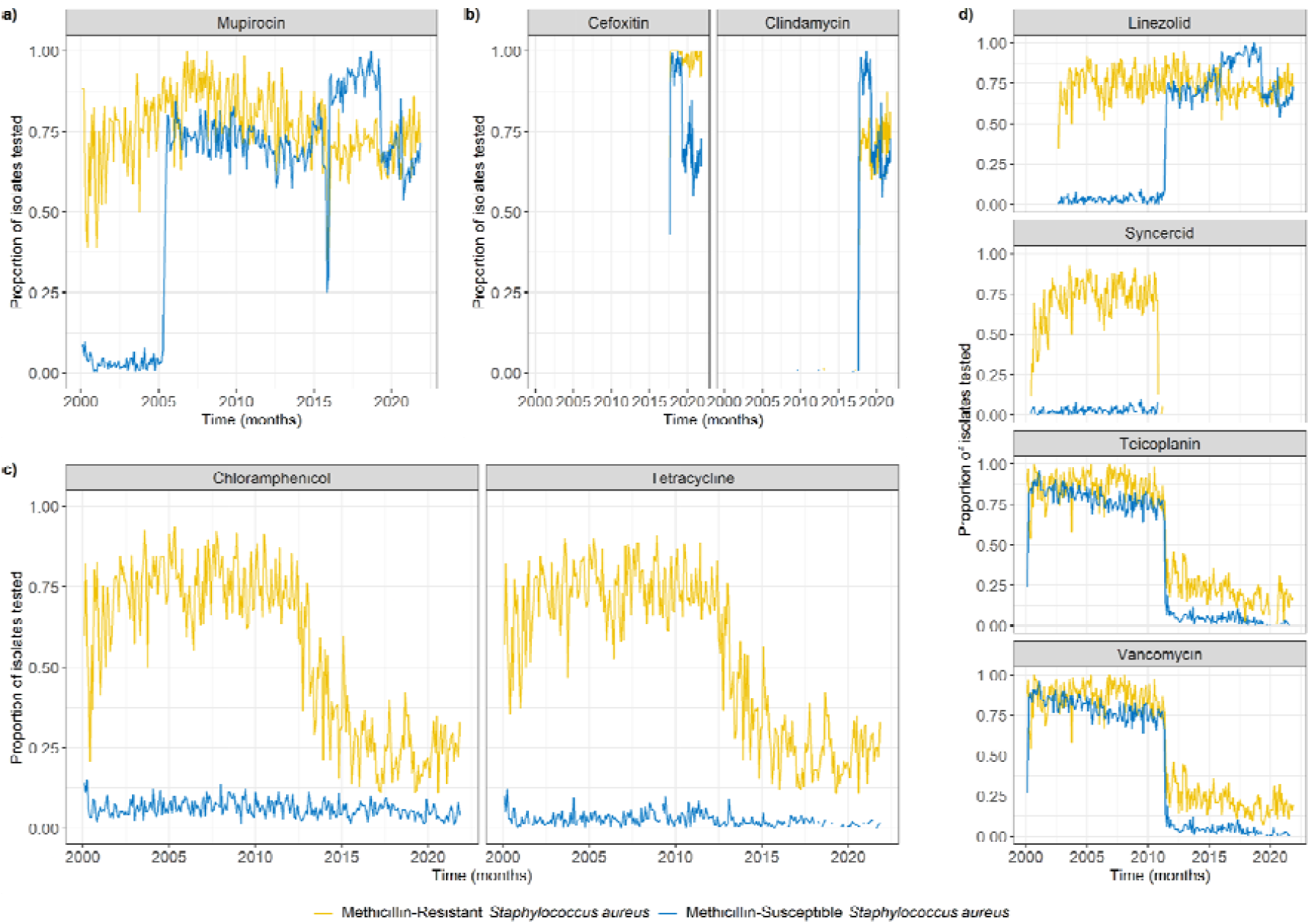
Change in proportion of *S. aureus* isolates tested for different antibiotic susceptibilities over time. These were subjectively grouped in four categories to facilitate visualisation. **a) Antibiotic with a change in susceptibility testing potentially responsible for a change in resistance detected. b) Antibiotics for which susceptibility testing began in 2017. c) Antibiotics with a decrease in susceptibility testing in MRSA. d) Antibiotics with aligned changes in susceptibility testing in 2011**. Missing lines indicate that no susceptibility testing for the corresponding antibiotic was conducted at that time.

The proportions of MRSA isolates resistant to amikacin (alone or jointly with flucloxacillin), gentamicin (alone or jointly with ciprofloxacin), and rifampicin were correlated, with simultaneous peaks in 2005 and 2007 (Figure 3a, Supplementary Figure 4). For both MRSA and MSSA isolates, ciprofloxacin resistance showed an overall decreasing trend since 2007 (more pronounced for MRSA), mupirocin resistance sharply decreased in 2004-2006, and clindamycin resistance has been increasing since 2017 (Figure 3b). Erythromycin resistance shows a decreasing trend for MRSA isolates, but an increasing one for MSSA isolates (Figure 3c). Trimethoprim resistance sharply declined for MSSA isolates in 2002, but remained stable for MRSA isolates (Figure 3c).

Mupirocin was the only antibiotic for which we could see a probable link between the proportion of isolates tested, and the proportion resistant. Mupirocin testing was always common in MRSA, but only became common for MSSA in 2007 (Figure 4a), which may explain the sharp decrease in the proportion of MSSA isolates found to be mupirocin-resistant in 2007 (Figure 3b).

Clindamycin and cefoxitin testing began in 2017, alongside more consistent cotrimoxazole testing (Figure 4b). Chloramphenicol and tetracycline testing were always at low levels for MSSA isolates, but decreased for MRSA isolates from 75% to 25% between 2012 and 2015 (Figure 4c).

Linezolid testing began in 2003, but was mostly reserved for MRSA isolates until 2011, after which the proportion of MSSA isolates tested for linezolid increased to the level of MRSA isolates (Figure 4d). Interestingly, this increase in 2011 aligned with a decrease in testing for syncercid (quinupristin and dalfopristin), teicoplanin and vancomycin, which affected both MRSA and MSSA isolates (Figure 4d) and was likely due to no resistance being seen in *S. aureus* isolates prior to 2011 (Supplementary Figure 6). These decreases, combined with the decrease in chloramphenicol and tetracycline mentioned above, explained the 2011 decrease in median number of tested resistances for MRSA isolates seen in Figure 2a.

### Dynamics of patient-level *S. aureus* AMR diversity

10,128/22,206 (45.6%) patients had more than one isolate recorded, allowing us to explore MRSA / MSSA and then further antibiogram diversity (Figure 5).

**Figure 5:**
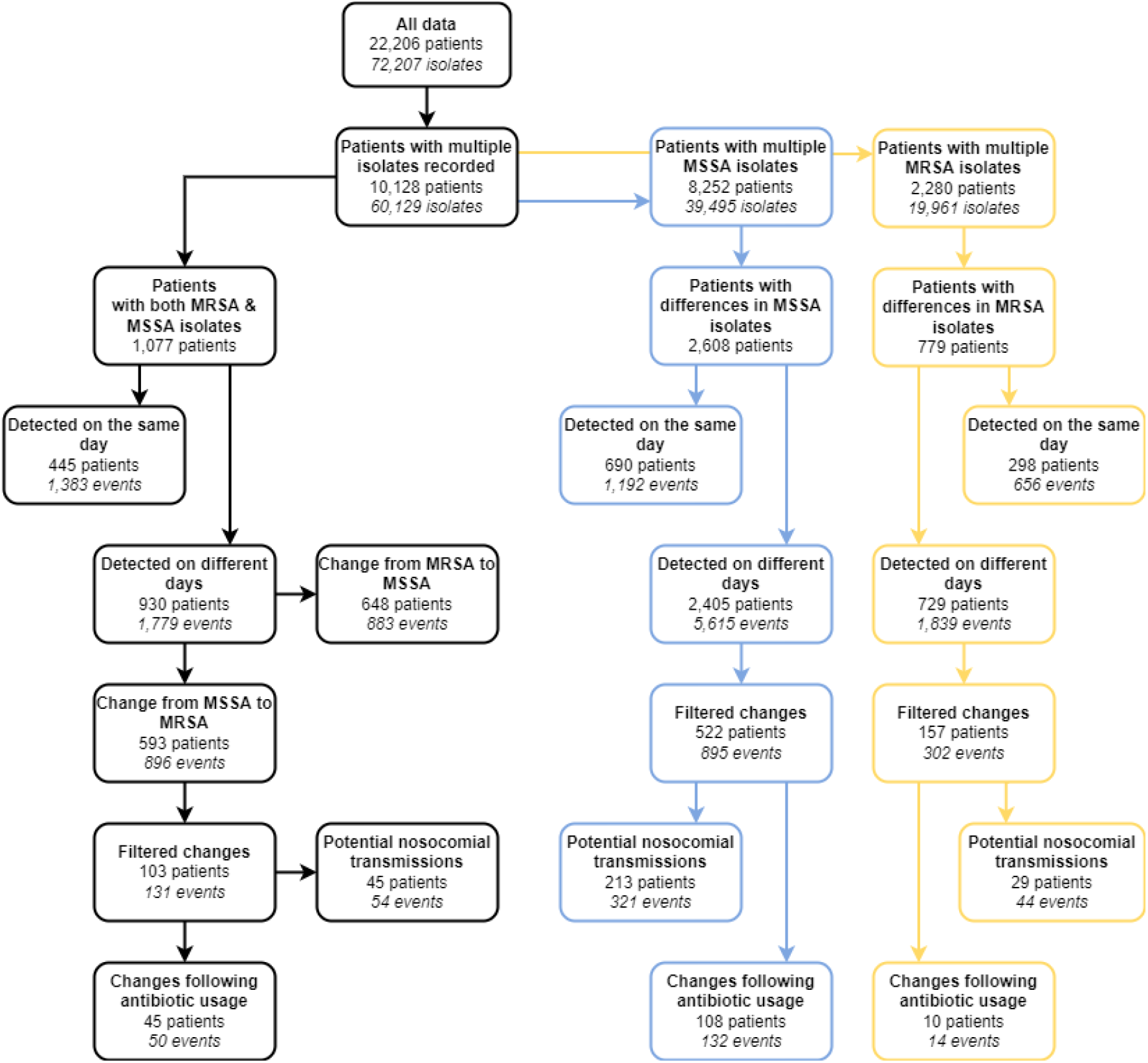
Data filtering process to identify patient-level AMR phenotypic diversity. Arrows indicate narrowing of the data for desired characteristics. Filtering for meticillin-susceptible *S. aureus* (MSSA) is shown in blue, and in yellow for meticillin-resistant (MRSA). A difference in MSSA or MRSA isolates corresponds to at least one difference in the antibiograms of these isolates. Categories may not be mutually exclusive (e.g. a patient could have both multiple MRSA isolates and multiple MSSA isolates recorded). Filtered changes: changes where both isolates were sampled during a single hospitalisation period, with at least 3 days between the isolates.

#### MRSA and MSSA diversity

We identified 1,077/22,206 (4.9%) patients for which both MRSA and MSSA isolates have been reported at any point in time. In 445 of these patients (41%, 2.0% of all patients), MRSA and MSSA isolates were detected on the same day, and in most cases (84%) from the same source (e.g. nose). The proportion of all patients positive for *S. aureus* simultaneously positive for MRSA and MSSA varied annually between 0.038 (2002) and 0.008 (2014) (Figure 6a).

**Figure 6:**
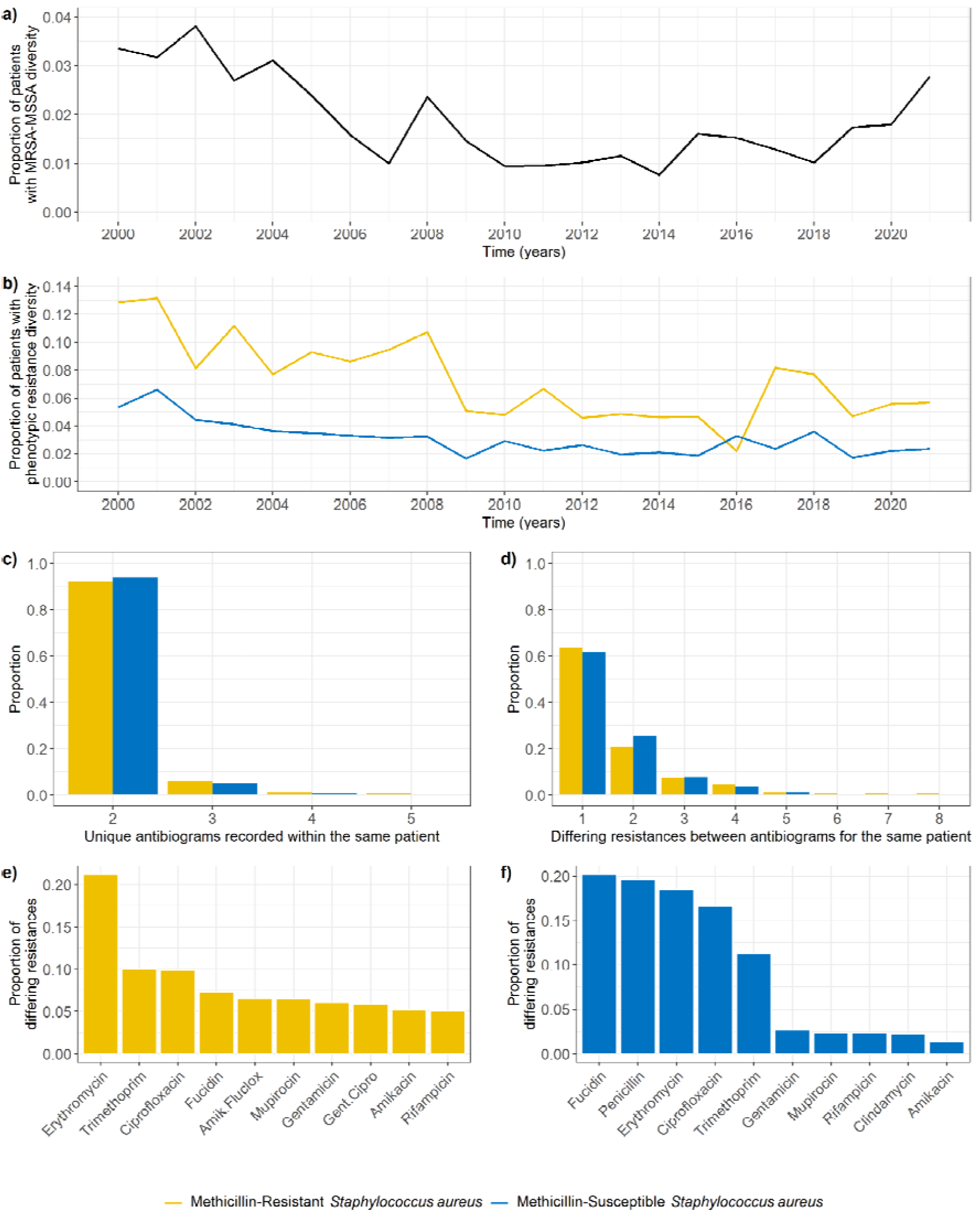
Patient-level *S. aureus* phenotypic AMR diversity on the same day. a) Annual proportion of patients for which both MRSA and MSSA isolates were detected on the same day, out of all patients positive for *S. aureus* in that same period. b) Annual proportion of patients for which diverse MRSA (yellow) or MSSA (blue) populations were detected on the same day, out of all patients positive for MRSA or MSSA in that same period. c) Number of unique antibiograms recorded for patients with diverse MRSA or MSSA populations on the same day. d) Number of differing resistances between antibiograms recorded for patients with diverse MRSA or MSSA populations on the same day. e) Most common differing resistances between MRSA isolates detected within the same patient on the same day. f) Most common differing resistances between MSSA isolates detected within the same patient on the same day.

#### Detected phenotypic resistance diversity

950 unique patients (4.3% of all patients) had multiple unique MSSA (690 patients) or MRSA (298 patients) antibiograms recorded on the same day (Figure 5). The isolates with different antibiograms were generally sampled from the same source (64%).

The percentage of patients with phenotypic resistance diversity was greater for patients with MRSA than for patients with MSSA (Figure 6b). In 90% of instances where diversity was found, the number of unique antibiograms recorded was 2 (Figure 6c), and in 90% of instances less than 4 resistances differed between the antibiograms (Figure 6d). Surprisingly, these values did not substantially differ between MRSA and MSSA populations (Figure 6c-d). The most common differing resistance between MRSA isolates was erythromycin, whilst other differences were homogeneously distributed amongst several antibiotics (Figure 6e). On the other hand, differences in MSSA isolates occurred predominantly in five antibiotics: fucidin, penicillin, erythromycin, ciprofloxacin, and trimethoprim (Figure 6f).

### Mechanisms driving patient-level dynamics of *S. aureus* AMR diversity

#### Dynamics of MRSA acquisition

In the 1,077 patients with both MRSA and MSSA isolates we detected 883/896 events (648/593 patients) where an MRSA/MSSA isolate was first detected, later followed by an MSSA/MRSA isolate respectively (Figure 5).

Focusing on the more clinically worrying events of a change from MSSA to MRSA, we found 131 events (103 patients) which occurred within single hospitalisations. The median delay between the detection of the MSSA and MRSA isolates was 9 days (Figure 7a). As a comparison, the distribution of delays between any two subsequent isolates recorded for patients in a single hospitalisation (excluding delays shorter than 2 days) had a heavier tail (Figure 7b). Patients in this group had substantially higher lengths of stay (median: 76 days, interquartile range: 28.25-239.75) compared to the lengths of stay of all patients in our dataset (Supplementary Figure 8, median: 7 days, IQR: 4-15, excluding stays shorter than 2 days).

**Figure 7:**
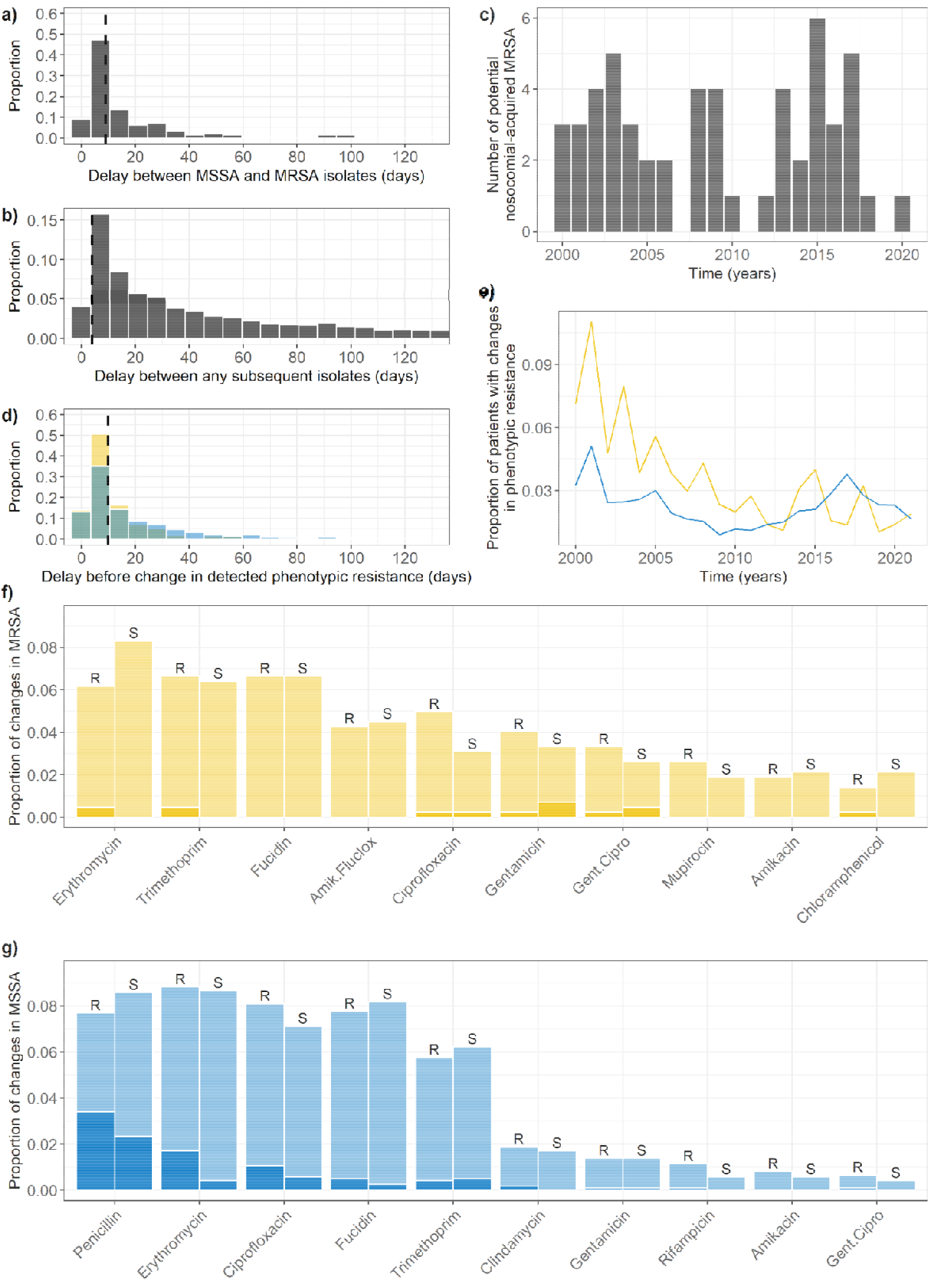
Changes in patient-level *S. aureus* phenotypic diversity over time. **a) Delay between detection of an MSSA and MRSA isolate in patients where the change happened within a single hospitalisation period.** The dashed line shows the median delay (9 days). Bin size is 7 days. Delays smaller than 2 days were not included. **b) Delay between detection of any two subsequent isolates in patients**. The dashed line shows the median delay (4 days). Bin size is 7 days. Delays smaller than 2 days were not included. We excluded 8,744 delays (32%) greater than 130 days from the figure for ease of comparison. **c) Potential MRSA nosocomial transmission events**. These are events from panel a) where an MRSA isolate was identified in another patient in the same ward immediately before the change. **d) Delay between detection of two isolates with a different antibiogram**. The dashed line shows the median delay (8 days). Bin size is 7 days. Delays smaller than 2 days were not included. We excluded 35 delays greater than 130 days from the figure. **e) Proportion of patients with differences in the antibiograms of their subsequent isolates over time**. MSSA and MRSA isolates are separated. **f) Most common changes detected in MRSA populations in single patients. g) Most common changes detected in MSSA populations in single patients**. Proportion of changes shown by antibiotic, type of change (R: susceptible to resistant, S: resistant to susceptible), and exposure to an antibiotic of the same class anytime between the original isolate, and the changed isolate (transparent: no, solid: yes). Amik.fluclox: joint amikacin and flucloxacillin resistance, gent.cipro: joint gentamicin and ciprofloxacin resistance.

We identified 50 changes from MSSA to MRSA in 45 patients which were preceded by any antibiotic use. Compared to events where the change was from MRSA to MSSA, we found no statistically significant link between the proportion of patients who were exposed to any antibiotic and the type of change after exposure (0.38 for MSSA to MRSA events, 0.37 for MRSA to MSSA; *X*^2^ = 0.02, df = 1, *p* = 0.90). We also identified 54 potential nosocomial acquisitions of MRSA, in 45 patients, with incidence varying over the years (Figure 7c).

52 detections of MRSA after MSSA in a single hospitalisation (in 44 patients) could not be linked to MRSA detected in the same ward or antibiotic exposure.

#### Dynamics of phenotypic resistance diversity

Of the 1,197 phenotypic resistance diversity events in 659 unique patients (3.0% of all patients, Figure 5), more were in MSSA isolates (895 events in 522 patients) than MRSA isolates (302 events in 157 patients). However, these events occurred relatively more often in patients who ever tested positive for MRSA (4.5%) than in patients who ever tested positive for MSSA (2.6%). The median delay between differing antibiograms was 10 days (Figure 7d). These events occurred more frequently in patients with MRSA than with MSSA, although this varied over the years (Figure 7e). In 30% of events, the identification of a new subpopulation in a patient was immediately preceded by this patient being in the same ward as another patient positive for this subpopulation (Figure 5).

We found some overlap between the changes in resistances most commonly detected (Figure 7f-g) and the antibiotic susceptibility tests most commonly conducted (Figure 2c-d). These changes almost equally represented gains and losses of resistance, regardless of the antibiotic (Figure 7f-g). In total, only 12% of all the changes we detected were preceded by patient exposure to an antibiotic of the same class as that of the change in the interval between the two differing antibiograms (Figure 7f-g). Interestingly, we noted several changes from resistance to susceptibility following antibiotic exposure (Figure 7f-g). The most common antibiotic resistance changes did not align with the most common antibiotic exposures in patients (Supplementary Figure 9).

We found no evidence for a causative link between antibiotic exposure and any change in resistance, as the proportion of all patients exposed to antibiotics who had a change detected (0.09) was slightly lower than those who were not exposed to antibiotics and still had a change detected (0.11; *X*^2^ = 6.70, degrees of freedom = 1, *p* < 0.01). Resistance genes for all antibiotics where changes were most commonly detected can be found on MGEs in *S. aureus* [44], except for ciprofloxacin and rifampicin, which occur mostly via mutations [6].

Overall, we identified 776 events (65%) where a change in phenotypic diversity occurred without previous exposure to antibiotics, nor apparent between-patient transmission.

## Discussion

### Results summary

Analysis of over 70,000 isolates collected routinely from 20,000 patients over 20 years at Great Ormond Street Hospital (GOSH) revealed substantial changes in resistance proportions at the hospital level. Testing regimens also varied, emphasising the importance of understanding screening strategies to study these trends. For example, mupirocin resistance changes could be linked to screening changes reflecting a close link between local context knowledge (treatment failure monitoring) and screening. Substantial declines in fluoroquinolone resistance, previously unnoted, are likely linked to national guidelines instructing a change in use, suggesting AMR can be reduced by such interventions.

This data also revealed that patient-level *S. aureus* populations are frequently diverse, with patients carrying heterogeneous populations of both MRSA and MSSA bacteria. We equally observed both gains and losses in phenotypic resistances of isolates over time at the patient level, suggesting considerable within-host diversity and evolution. The majority of patients with changes in detected resistance within *S. aureus* could not have this AMR evolution linked easily to detected nosocomial transmission or antibiotic exposure. Unexpectedly, antibiotic exposure was equally linked to a loss or acquisition of phenotypic resistance.

### Hospital-level AMR diversity

Analysing 20 years of data allowed us to reveal long term changes in testing (tetracycline / teicoplanin) and resistance (erythromycin) that would be missed in shorter scale analysis. Notably, the proportion of isolates resistant to mupirocin decreased as a consequence of a substantial increase in the proportion of isolates tested for susceptibility to this antibiotic. This is a well-known type of bias where conducting tests in a group consisting largely of severely ill patients leads to an overestimation of the prevalence of resistant isolates [45].

The decrease in ciprofloxacin resistance likely demonstrates that policies to reduce antibiotic usage can successfully lead to a decrease in resistance over time. In 2006, a UK-wide policy was introduced to control *Clostridium difficile* infections by limiting fluoroquinolone usage in all hospitals [46,47]. This aligns with the start of the decrease in ciprofloxacin resistance measured in our *S. aureus* isolates, particularly for MRSA isolates as 90% were resistant to ciprofloxacin before 2007, down to 40% in 2021 (Figure 3). This association could be confirmed by obtaining antibiotic usage data from all patients at GOSH, which we did not have access to in this study.

We also noted amikacin, gentamicin and rifampicin as examples of rare but highly correlated resistances. To the best of our knowledge, although joint amikacin and gentamicin resistance is well-known as both are aminoglycoside antibiotics [48,49], the biological mechanism explaining the correlation with rifampicin resistance is unknown.

A subset of isolates were CC typed over time, indicating that CC22 was the dominant clone prior to 2011 which is typical for the UK [23]. Post-2011, typing was extended to suspected PVL-positive MRSA and MSSA, and the dominant clones identified reflect the expected CC types.

Previous studies have highlighted the importance of imported MRSA from abroad [21]. A link may exist between the increase in MRSA isolates between 2014 and 2020, and the ethnicity of patients admitted at GOSH. We did find that changes in the proportion of patients with an ethnicity other than “White British” were aligned with changes in the proportion of *S. aureus* isolates which were MRSA, suggesting that this link does exist in our data. In addition, since patients from outside the UK were no longer admitted to GOSH during the first COVID-19 lockdown in March 2020, this may also explain the decrease in MRSA isolates seen at that time.

The multiple changes in testing strategy we identified in 2011 were linked to a change in the head of infection prevention and control. Following identification of MRSA in a patient, it is common to swab multiple other potential colonisation sites to identify the presence of *S. aureus*. However, any isolate subsequently identified through these additional swabs would not undergo complete antibiotic susceptibility testing. This explains the higher number of isolates since 2011 with fewer antibiotic susceptibility testing conducted, as well as the 10,029 isolates in our data with no antibiotic susceptibility results.

### Patient-level AMR diversity

In single patients with *S. aureus* AMR diversity, MRSA populations were on average more diverse than MSSA populations. This could be explained by more MRSA samples being taken per patient than MSSA, consistent with UK testing guidelines, and hence increasing the probability to detect diverse subpopulations [36]. However, this prevalence of diversity changed over time while the number of isolates recorded per patient did not, suggesting there are other unknown factors affecting diversity. Surprisingly, when diversity was detected, the number of differences was similar regardless of whether they occurred in MSSA or MRSA populations (two unique antibiograms detected, with less than four differences between antibiograms). As MRSA isolates carry more resistances, we expected to see more instances where diversity was detected than in MSSA, and a greater amount of diversity (we did not see the latter). This could firstly be due to detection limits, as sampling in the diagnostic laboratory relying on plating may not be powerful enough to detect profiles representing only small proportions of the total *S. aureus* population [11]. Alternatively, a previous study of *S. aureus* diversity in piglets found evidence of simultaneously high rates of gain and loss of MGEs in *S. aureus*, which may explain why less diversity is detected here, as some subpopulations of *S. aureus* carrying different resistance genes may only be present at small proportions and therefore undetected [32].

Our estimates for diversity are lower than those reported in previous studies. Whilst we report 2.0% of all patients ever positive for *S. aureus* carried both MSSA and MRSA simultaneously at some point, a previous study estimated this value to be 21% in children sampled before surgery [12]. This same study identified multiple *S. aureus* genotypes in 30% of hospital patients, while our estimate of diversity (defined as detection of multiple MSSA or MRSA isolates with different antibiograms on the same day, in a patient with at least one MSSA or MRSA isolate detected) was 3.5% for patients with MSSA detected, and 8.5% for patients with MRSA detected. On the other hand, a second study found that 6.6% of individuals colonised by *S. aureus* carried more than one strain, defined using pulsed field gel electrophoresis [13]. These differences may be partly due to different definitions of diversity across studies, and the fact that these previous studies collected data specifically to identify diversity, whilst we used routinely collected data with reduced sampling. Finally, a study in patients colonised with MRSA found that 24% were colonised by more than one phenotypically distinct isolate [10]. Interestingly, this study identified a maximum of three distinct isolates in a single patient, and two in the remaining eight patients with diversity. This is consistent with our result that, in patients with diversity, we generally detected only two MSSA or MRSA subpopulations (only two unique antibiograms), although we were able to detect up to five unique antibiograms within a single patient on the same day (Figure 6c).

### Within-host evolution as a driver of AMR diversity

We identified 131 events (103 patients) which correspond to patients acquiring MRSA during their hospitalisation period. Of these events, 54 (41%, 45 patients) represent probable instances of nosocomial-acquired MRSA This likely represents a lower estimate of nosocomial-acquired MRSA, since we have not considered in this category the instances where patients initially tested negative for any *S. aureus* initially, and subsequently positive for MRSA. Note that we believe that the majority of MRSA carriers in the hospital would be detected as all are screened on entry for *S. aureus* and, if found, meticillin resistance.

There were 77 MRSA acquisition events (59%, 64 patients) where the patients did not share a ward with any other patient positive for MRSA immediately before the acquisition was reported. In such cases, the patients may have acquired MRSA from a source not monitored in our data, such as healthcare workers or environmental surfaces [7], or these events may represent incorrect detection. These events should be further investigated to clarify MRSA acquisition source. This emphasises our likely underestimate of nosocomial transmission.

We expected many changes in diversity due to acquired detected resistance to be linked to antibiotic exposure, whereby potentially undetected minority subpopulations are selected for. However, in the identified 1,197 events (3% of all patients) where a change in at least one phenotypic resistance was detected within a single hospitalisation period we found evidence supporting this for only 12% of all changes, and we did not find that patients with a change in resistance were significantly more likely to have been exposed to antibiotics of the same class in hospital, than patients with no change. Furthermore, some detected losses in resistance counterintuitively occurred following exposure to antibiotics belonging to the same class. Since some resistances are co-located on plasmids, antibiotic selection for one resistance may have incidentally selected for other resistances present on the same plasmid [44], but we could not estimate this here due to lack of genetic data.

A further 367 events (30%) could be linked to nosocomial transmission where detected changes in phenotypic resistances occurred after patients shared a ward with another patient for which an isolate with the same antibiogram had previously been detected. Movement of single MGEs carrying antibiotic resistance between patients is possible, but a previous study which measured this using detailed genomic data from more than 2,000 isolates of various bacteria species over 18 months only found a single instance where this may have occurred [50].

Hence, the majority of changes in phenotypic diversity (776 events, 65%) could not be explained by either antibiotic exposure or between-patient transmission. Therefore a third explanation could be within-host evolution via frequent gain and loss of antibiotic resistance genes in *S. aureus* populations. This was previously seen *in vivo* in gnotobiotic piglets, and could explain changes for some resistances which are known to be located on MGEs [32].

However, we also saw changes in resistance to ciprofloxacin and rifampicin, which are gained via mutations instead of acquisition of an MGE [6]. We also failed to see changes in resistances we might have expected to move frequently, such as clindamycin, tetracycline, and chloramphenicol, present on many MGEs in *S. aureus* [44]. This may indicate varying rates of gain and loss for different genes or fitness effects such as a high cost of resistance (e.g. for rifampicin [51]). The major mechanism for horizontal gene transfer in *S. aureus* is transduction by bacteriophage, with phage capable of transduction found in at least 50% of *S. aureus* within-host populations [10,52–54]. Previous work suggested that different genes may not be transferred at the same rate by this process, which may explain the variations we have seen here [32,54].

### Strengths and limitations

To the best of our knowledge, this is the first time that such a large dataset has been used to search for evidence of AMR diversity in *S. aureus*. A key strength of our analysis is its longitudinal nature, which allowed us to explore the dynamics of this process with colonising and invasive MRSA and MSSA isolates instead of observing only snapshots as multiple studies have previously done. However, as GOSH specialises in paediatric care, the patients in our analysis belong to a group which may not be representative of the entire population: *S. aureus* infections are typically less severe in children than adults [55] and colonisation may be more prevalent in children [56]though the proportion of children with MRSA versus MSSA is likely similar as in adults [57].

Phenotypic resistances, which we focused on here, are likely more clinically meaningful to compare than the presence or absence of resistance genes, as previous studies have done [10], because genotypic traits of antibiotic resistance may not always translate to phenotypic resistance [58]. However, measured diversity in phenotypic resistance is lower than genotypic, as bacteria of the same species displaying resistance to the same antibiotics will be considered identical, even if the resistance genes they carry are different. The fact that we are still able to see evidence of AMR diversity in *S. aureus* populations using phenotypic data only, strongly supports the importance of this diversity, and that it is likely to have consequences on the estimates of the health burden of infections and AMR evolution

The major limitation of our work is that all the antibiotic susceptibilities were determined from routine clinically-motivated samples. The sampling strategy was not designed to fully capture diversity in *S. aureus* populations within single patients, which may explain our lower estimates compared to previous studies specifically designed for this purpose. Although multiple colonies are separately subcultured only when they are visually different on the plate (e.g. different sizes or colours) [59], this process has never been audited, and generally only 2-3 colonies are selected for subculture and antibiotic susceptibility testing. Previous work has shown that different strains may coexist at various proportions within a single *S. aureus* population, and that strains in minority may be missed if less than 18 colonies are sampled [10]. Crucially, this means that some of the changes we report may simply represent the variability of sampling in the laboratory. Although we tried to account for this by restricting our analysis to changes in samples collected at least 3 days apart, this limitation is an inevitable consequence of our attempt to use routinely collected diagnostic microbiology laboratory data.

In addition, not all isolates are tested for all antibiotics and we did not consider that a change from no test to resistant or susceptible was a valid change in resistance, which means that we may potentially have missed some changes. However, this limitation is less relevant for our results on MSSA-MRSA diversity due to the policy of screening all patients upon admission to the hospital for *S. aureus* colonisation, and systematic and accurate testing for MRSA.

Susceptibility testing in the diagnostic microbiology laboratory is conducted using disk diffusion or gradient methods. As these methods rely on breakpoints to classify isolates as susceptible or resistant, there may be some subjectivity in the classification. However, through discussions with clinical microbiologists at GOSH, we confirmed that all the antibiotics were equally likely to be affected by this subjectivity, meaning this potential bias is homogeneously distributed in our data.

In any case, if the sampling limitations listed above are actually responsible for all the changes in phenotypic resistances we detected, that would clearly indicate that the testing strategy in diagnostics laboratories frequently misses resistant subpopulations in patient samples. This would have important implications for treatment, as failure to identify resistance may lead to inappropriate antibiotic choices, treatment failures, and worse health outcomes for patients.

### Implications and next steps

We have shown here the value of routinely collected data both at the hospital level, to understand how hospital policies can affect AMR over time, but also at the patient level, to reveal potential drivers of AMR diversity. This was made possible through the framework developed by GOSH to store and make this data easily accessible for research purposes. Other healthcare institutions should develop systems like the GOSH DRIVE to record and analyse routinely collected data [60], as this may benefit these institutions directly, and allow new analyses to further improve our understanding of AMR. Improved sharing of information on data collection to accompany the public release of these routinely collected datasets will be beneficial to the scientific community more broadly.

Our results also illustrate the limitations of routinely collected hospital data, showing for example that some changes in antibiotic resistance detected over time may not correspond to true epidemiological trends, but rather are linked to changes in testing strategy. As this type of data is frequently used in secondary analyses to derive epidemiological trends in AMR prevalence, future studies should bear this important limitation in mind. Unless the microbiology and infectious disease context is clearly understood, the data is uninterpretable, hence clinical involvement is paramount. For example, here a lack of vancomycin resistance in *S. aureus* past 2011 is likely to have led to the decision to stop testing all MRSA isolates. This pragmaticism is logical but could lead to data misinterpretation and potential delayed detection of resistance arising. However, the former would also be detected through treatment failure. Ongoing collaborative discussions with hospital staff should be encouraged in any analysis of routine data, as they may provide explanations for unexpected changes in reported rates of AMR at the local level.

Repeating this work in other hospitals would provide valuable insights as to drivers of infection and resistance since *S. aureus* incidence can vary substantially within and between countries, and population structures such as dominant lineages vary geographically [25]. In particular, extending into adult populations and comparing hospitals with overlapping patient networks would improve generalisability, understanding how such diversity may be transmitted across hospital networks, as well as expanding to other key clinical pathogens [61–65].

Moreover, further studies with longitudinal sampling of patients, including both genotypic and phenotypic AMR diversity would improve our understanding of AMR evolution and movement, as well as patient outcomes. In addition, linked to the importance of transduction in *S. aureus* testing patient samples for the presence of phage, as was done previously at a small scale [10], would aid in determining the importance of transmission vs. selection vs. within-host movement of resistance genes.

## Conclusion

Our analysis of routinely collected data has revealed a complexity in resistance prevalence and sampling that needs emphasising as we move forward with estimating AMR burden and evolution. These nuances need to be recognised in any analysis of routine surveillance data with the pragmatism of a long-term reality of non-universal infrequent sampling, even in high income settings, meaning that the AMR community needs to work on developing methods that can account for and link patterns across multiple levels. Surprisingly we could not find clear evidence of antibiotic usage leading to resistance at the patient level, but did find evidence of a wider policy of reduced use of fluoroquinolones leading to a hospital wide decrease in resistance and emphasis the likely importance of within host variation and rapid shuffling of resistance genes in *S. aureus*.

## Data Availability

The raw datasets are the property of Great Ormond Street Hospital (GOSH) and cannot be shared publicly. The processed anonymised datasets and the code to run the analyses are publicly available in a GitHub repository (https://github.com/qleclerc/gosh_mrsa).

## Ethical approval

Ethical approval for this study was obtained both from Great Ormond Street Hospital (under ethical approval 17/LO/0008 for use of routine GOSH data for research), and the London School of Hygiene & Tropical Medicine (reference 26692).

## Author contributions

Conceptualization: QJL, LG, GMK; Data curation: QJL; Formal Analysis: QJL; Methodology: QJL, LG, GMK; Software: QJL; Supervision: JAL, LG, GMK; Validation: AC, HD, JH; Visualisation: QJL, JAL, LG, GMK; Writing - original draft: QJL; Writing - review & editing: QJL, AC, HD, JH, JAL, LG, GMK

## Acknowledgments

This work was facilitated by the GOSH Data Research, Innovation and Virtual Environments (DRIVE) unit. In particular, we would like to thank Mohsin Shah and Timothy Best for their help with the data extraction process.

