## Supplementary Material for "Quantifying patient- and hospital-level antimicrobial resistance dynamics in *Staphylococcus aureus* from routinely collected data"

**Supplementary Table 1: Matching information for antibiotics and antibiotic classes.**

| Antibiotic | Class |
|---|---|
| Amikacin | Aminoglycoside |
| Gentamicin | Aminoglycoside |
| Tobramycin | Aminoglycoside |
| Rifaximin | Ansamycin |
| Bedaquiline | Bedaquiline |
| Ertapenem | Carbapenem |
| Imipenem | Carbapenem |
| Meropenem | Carbapenem |
| Cefalexin | Cephalosporin |
| Cefiderocol | Cephalosporin |
| Cefixime | Cephalosporin |
| Cefotaxime | Cephalosporin |
| Cefoxitin | Cephalosporin |
| Ceftazidime | Cephalosporin |
| Ceftolozane | Cephalosporin |
| Ceftriaxone | Cephalosporin |
| Cefuroxime | Cephalosporin |
| Chloramphenicol | Cephalosporin |
| Chlorhexidine | Chlorhexidine |
| Clofazimine | Clofazimine |
| Dapsone | Dapsone |
| Ethambutol | Ethambutol |
| Ciprofloxacin | Fluoroquinolone |
| Levofloxacin | Fluoroquinolone |
| Moxifloxacin | Fluoroquinolone |
| Ofloxacin | Fluoroquinolone |
| Fosfomycin | Fosfomycin |
| Daptomycin | Lipopeptide |
| Azithromycin | Macrolide |
| Clarithromycin | Macrolide |
| Erythromycin | Macrolide |
| Metronidazole | Metronidazole |
| Mitomycin | Mitomycin |
| Aztreonam | Monobactam |
| Mupirocin | Mupirocin |
| Nitrofurantoin | Nitrofuran |
| Linezolid | Oxazolidinone |
| Tedizolid | Oxazolidinone |
| Amoxicillin | Penicillin |
| Ampicillin | Penicillin |
| Benzylpenicillin | Penicillin |
| Co-Amoxiclav | Penicillin |
| Flucloxacillin | Penicillin |
| Phenoxymethylpenicillin | Penicillin |
| Piperacillin | Penicillin |
| Pivmecillinam | Penicillin |
| Temocillin | Penicillin |
| Colistin | Polypeptide |
| Protionamide | Protionamide |
| Pyrazinamide | Pyrazinamide |
| Rifampicin | Rifamycin |
| Co-Trimoxazole | Sulfonamide |
| Sulfadiazine | Sulfonamide |
| Trimethoprim | Sulfonamide |

| | | | |
|---|---|---|---|
| Fucidin | Fucidin | Doxycycline | Tetracycline |
| Teicoplanin | Glycopeptide | Minocycline | Tetracycline |
| Vancomycin | Glycopeptide | Tetracycline | Tetracycline |
| Isoniazid | Isoniazid | Tigecycline | Tetracycline |
| Clindamycin | Lincosamide | Thalidomide | Thalidomide |

**Supplementary Table 2: Patient characteristics.** IQR: interquartile range. LoS: length of stay. Hospital stays shorter than 2 days are not counted in the table and not included in the median LoS estimation.

| Year | Number of unique patients | Female patients (%) | Median age (IQR) | "Other Groups" ethnicity (%) | Number of hospital stays | Median LoS (IQR) |
|---|---|---|---|---|---|---|
| **2000** | 1103 | 44.35 | 4 (1-10) | 49.37 | 2311 | 5 (3-11) |
| **2001** | 1260 | 42.49 | 4 (1-10) | 50.25 | 2770 | 4 (3-10) |
| **2002** | 1286 | 43.49 | 4 (1-10) | 46.32 | 2836 | 4 (2-10) |
| **2003** | 1223 | 45.04 | 5 (1-10) | 46.58 | 2791 | 4 (2-10) |
| **2004** | 1384 | 44.03 | 5 (1-10) | 48.52 | 2875 | 4 (2-10) |
| **2005** | 1419 | 43.93 | 5 (1-10) | 47.02 | 2558 | 4 (2-10) |
| **2006** | 1451 | 42.14 | 5 (1-10) | 48.44 | 2520 | 4 (2-11) |
| **2007** | 1501 | 45.4 | 5 (1-11) | 48.72 | 2381 | 5 (2-11) |
| **2008** | 1481 | 45.61 | 4 (1-10) | 49.43 | 2409 | 5 (2-10) |
| **2009** | 1435 | 43.38 | 4 (1-10) | 49.61 | 2312 | 5 (2-11) |
| **2010** | 1587 | 41.51 | 4 (1-10) | 53.39 | 2413 | 5 (3-11) |
| **2011** | 1573 | 44.05 | 4 (1-10) | 49.24 | 2330 | 5 (2-11) |
| **2012** | 1478 | 45.23 | 4 (1-10) | 50.22 | 2308 | 5 (3-13) |
| **2013** | 1558 | 43.29 | 4 (1-10) | 51.80 | 2387 | 5 (3-13) |
| **2014** | 1569 | 44.52 | 5 (1-11) | 52.14 | 2254 | 5 (3-12) |
| **2015** | 1490 | 46.44 | 5 (1-10) | 54.41 | 2227 | 6 (3-13) |
| **2016** | 1246 | 45.86 | 5 (1-11) | 58.42 | 2222 | 6 (3-13) |
| **2017** | 1321 | 45.75 | 5 (1-11) | 57.52 | 2164 | 5.5 (3-14) |
| **2018** | 1280 | 45.06 | 5 (1-10) | 61.40 | 2265 | 6 (3-14) |
| **2019** | 1497 | 44.01 | 5 (1-10) | 57.61 | 2124 | 6 (3-14) |

| | | | | | | |
|---|---|---|---|---|---|---|
| **2020** | 1337 | 46.30 | 5 (1-11) | 58.19 | 1895 | 6 (3-14) |
| **2021** | 1366 | 45.61 | 6 (1-11) | 55.54 | 1720 | 6 (3-13) |

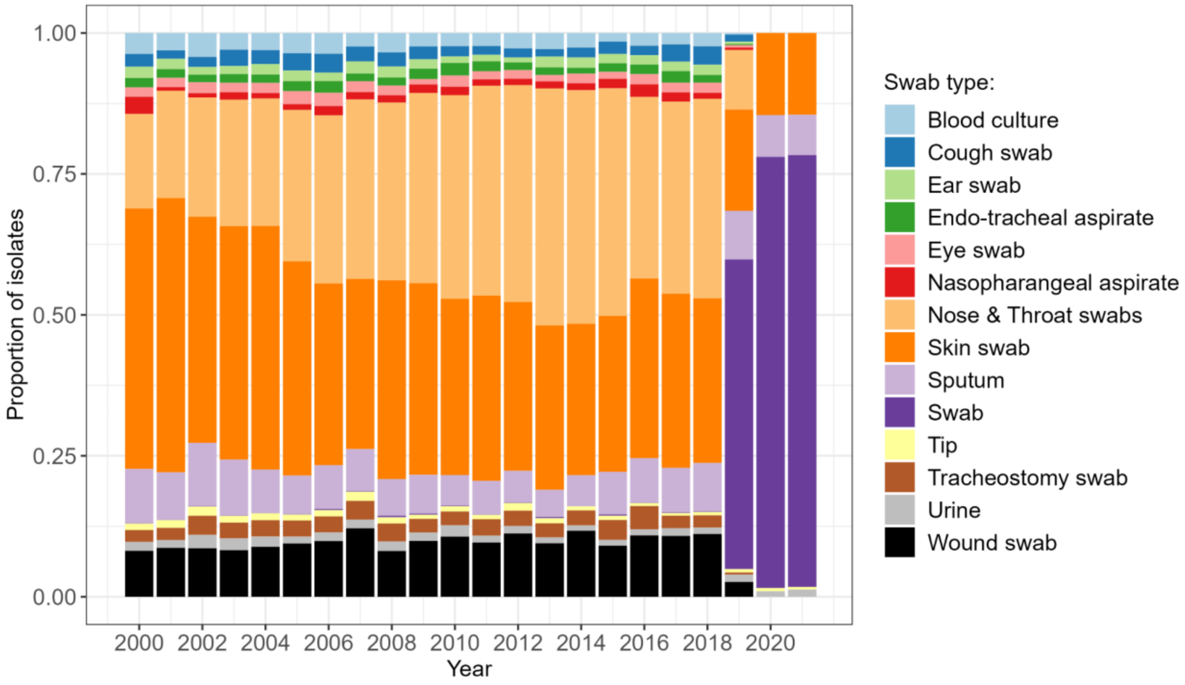


**Supplementary Figure 1: Distribution of *S. aureus* isolate sources over time.** Only the 14 most common sources across the entire dataset (covering 93% of all isolates) are shown for ease of visualisation.

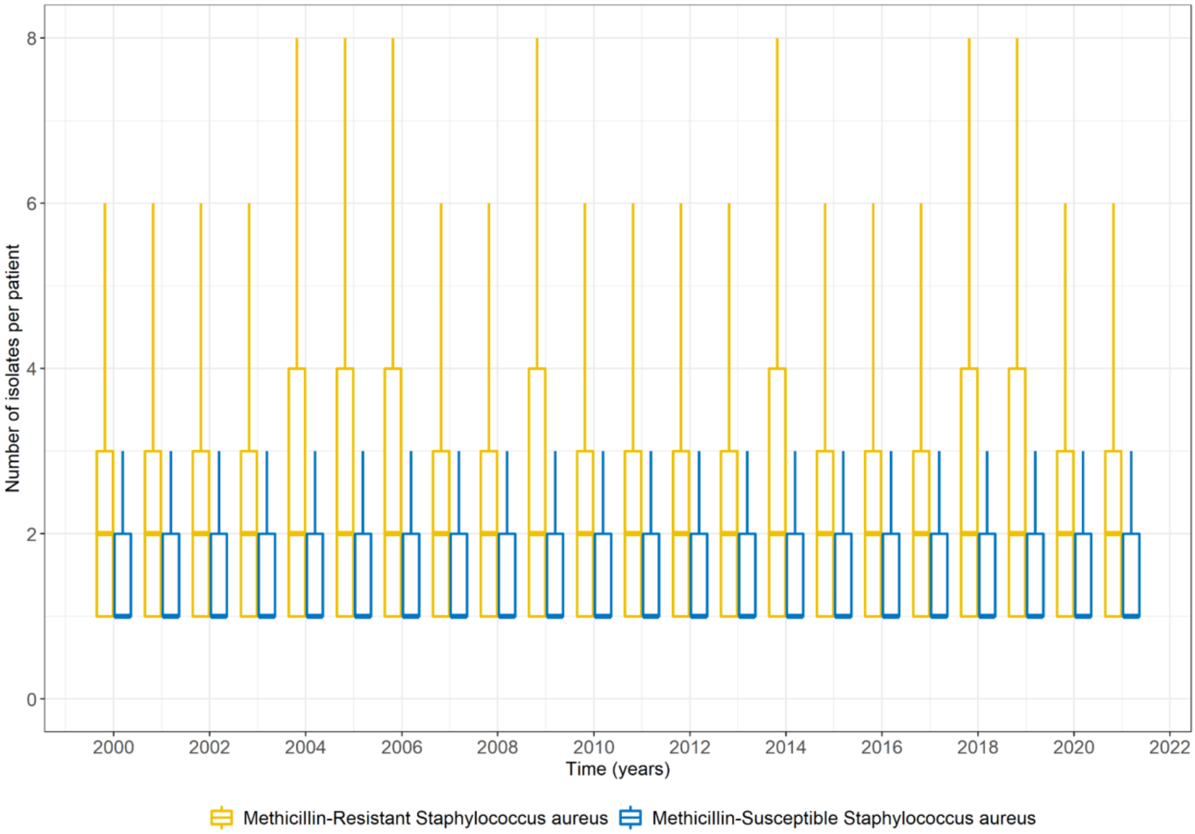


**Supplementary Figure 2: Number of MRSA or MSSA isolates per patient per year.** Bold lines show the median, box limits show the interquartile range, line limits show the largest value within 1.5 interquartile range above the 75th percentile and smallest value within 1.5 interquartile range below the 25th percentile. Outliers are not shown on the figure.

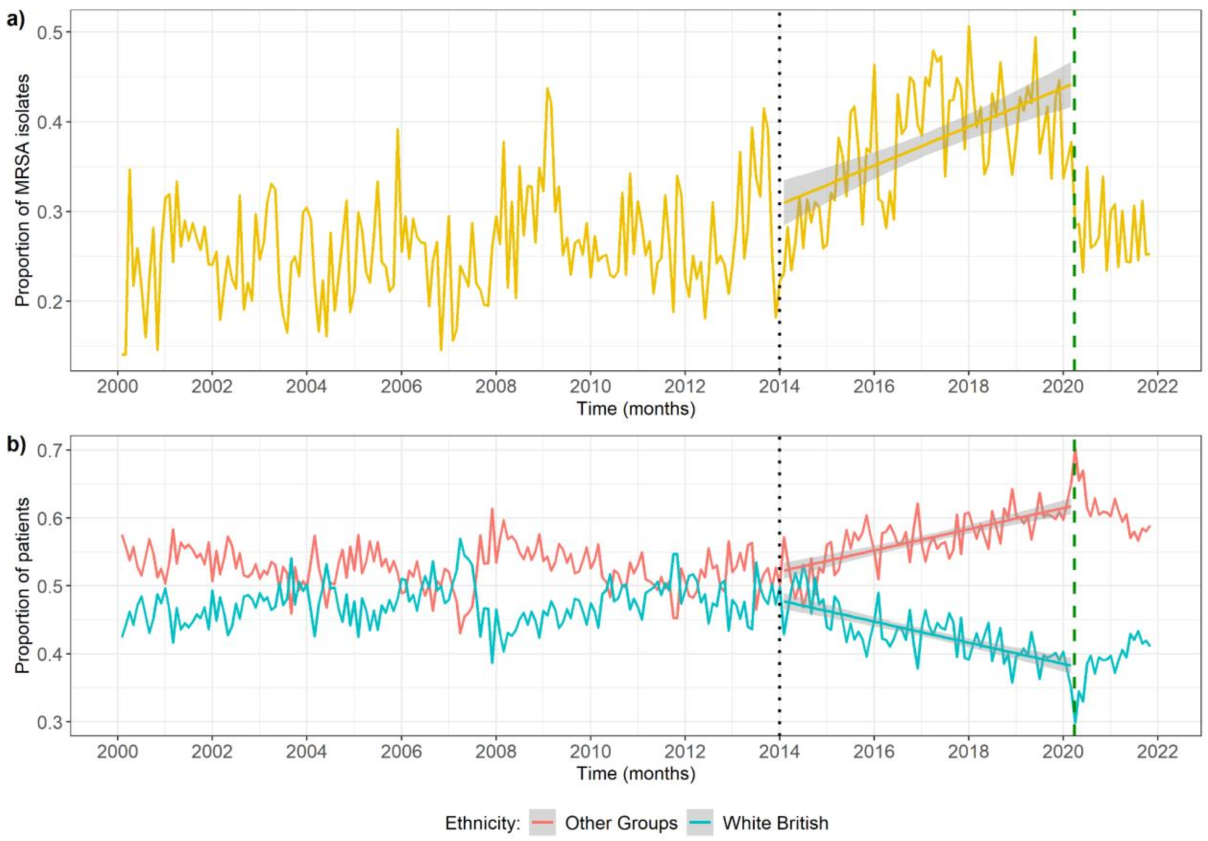


**Supplementary Figure 3: Proportion of all *S. aureus* isolates which are meticillin-resistant (a) and ethnicity of patients in our dataset (b).** Vertical green dashed line shows the date when lockdown began in the UK during the first wave of the COVID-19 pandemic (29/03/20). Patients with no recorded ethnicity are excluded from the analysis. The interval between the dotted black line (01/01/14) and the green line shows the period where the proportion of MRSA isolates and proportion of patients with an ethnicity of "Other Groups" both increased. The solid lines show the linear regression trendlines between time and these variables.

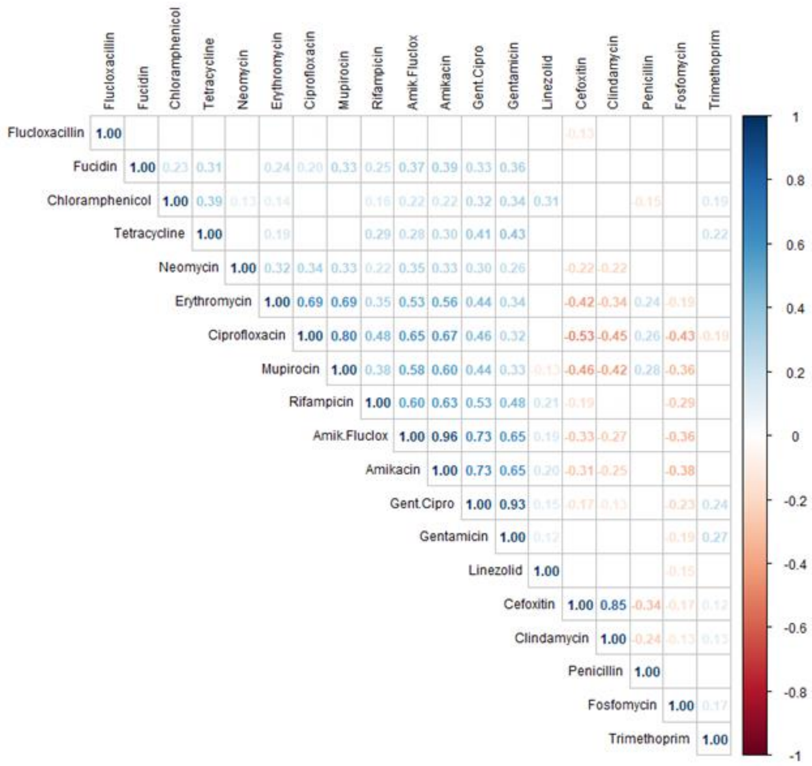

**Supplementary Figure 4: Spearman's correlation coefficients between proportions of meticillin-resistant *S. aureus* isolates resistant to different antibiotics over time.** Antibiotics included are those with at least 50 susceptibility tests conducted over the entire time period (2000-2021). Only significant coefficients are shown (p value < 0.05).

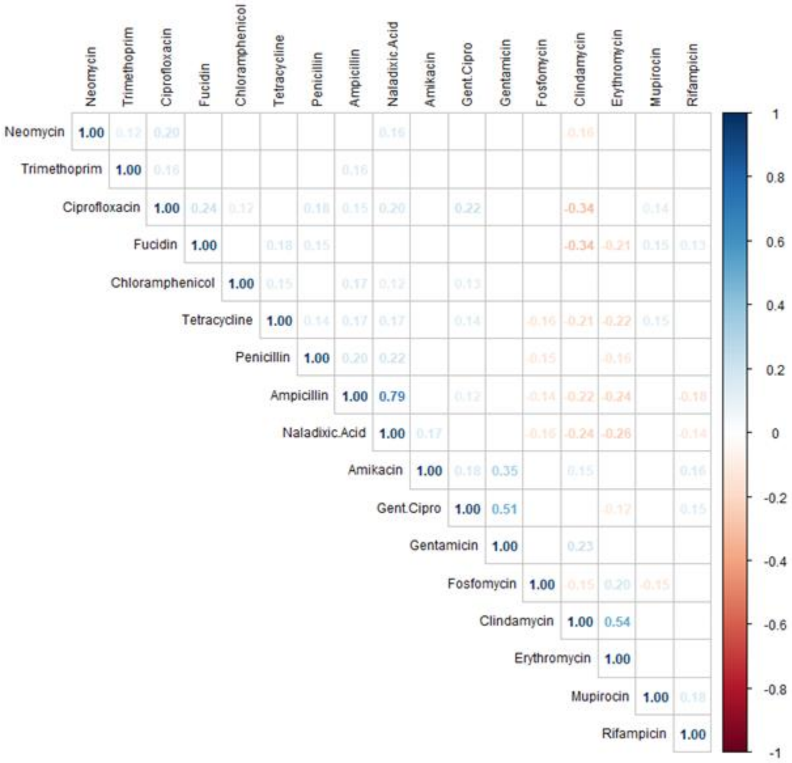


**Supplementary Figure 5: Spearman's correlation coefficients between proportions of meticillin-susceptible *S. aureus* isolates resistant to different antibiotics over time.** Antibiotics included are those with at least 50 susceptibility tests conducted over the entire time period (2000-2021). Only significant coefficients are shown (p value < 0.05).

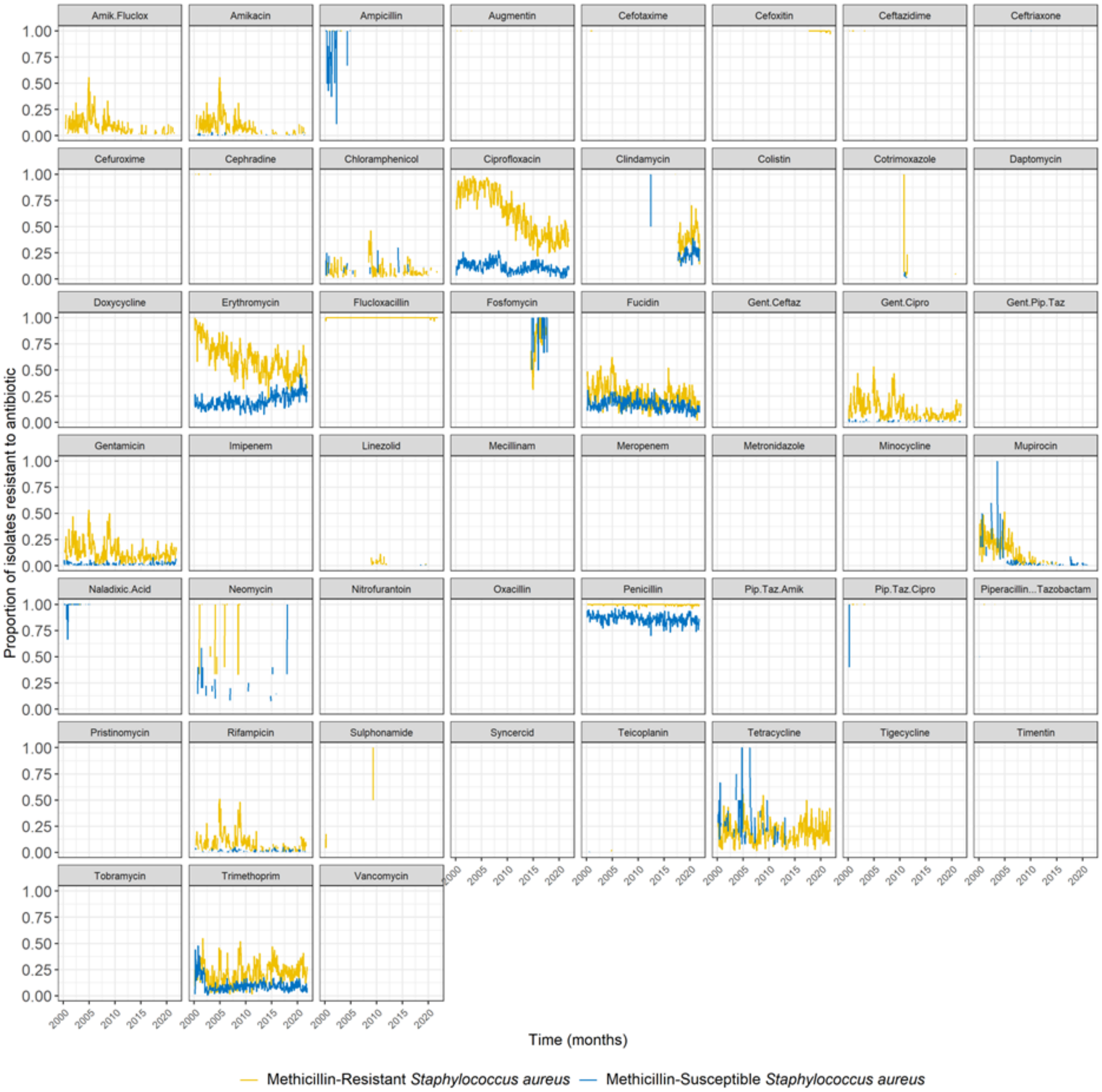


**Supplementary Figure 6: Change in proportion of *S. aureus* isolates resistant to antibiotics over time, out of those tested for resistance to the corresponding antibiotic.** Amik.Fluclox: joint amikacin and flucloxacillin resistance, Gent.Ceftaz: joint gentamicin and ceftazidime resistance, Gent.Cipro: joint gentamicin and ciprofloxacin resistance, Gent.Pip.Taz: joint gentamicin and piperacillin-tazobactam resistance, Pip.Taz.Amik: joint piperacillin-tazobactam and amikacin resistance, Pip.Taz.Cipro: joint piperacillin-tazobactam and ciprofloxacin resistance. Missing lines indicate that no resistance to the corresponding antibiotic was detected at that time (which may be due to no testing having been conducted at that time).

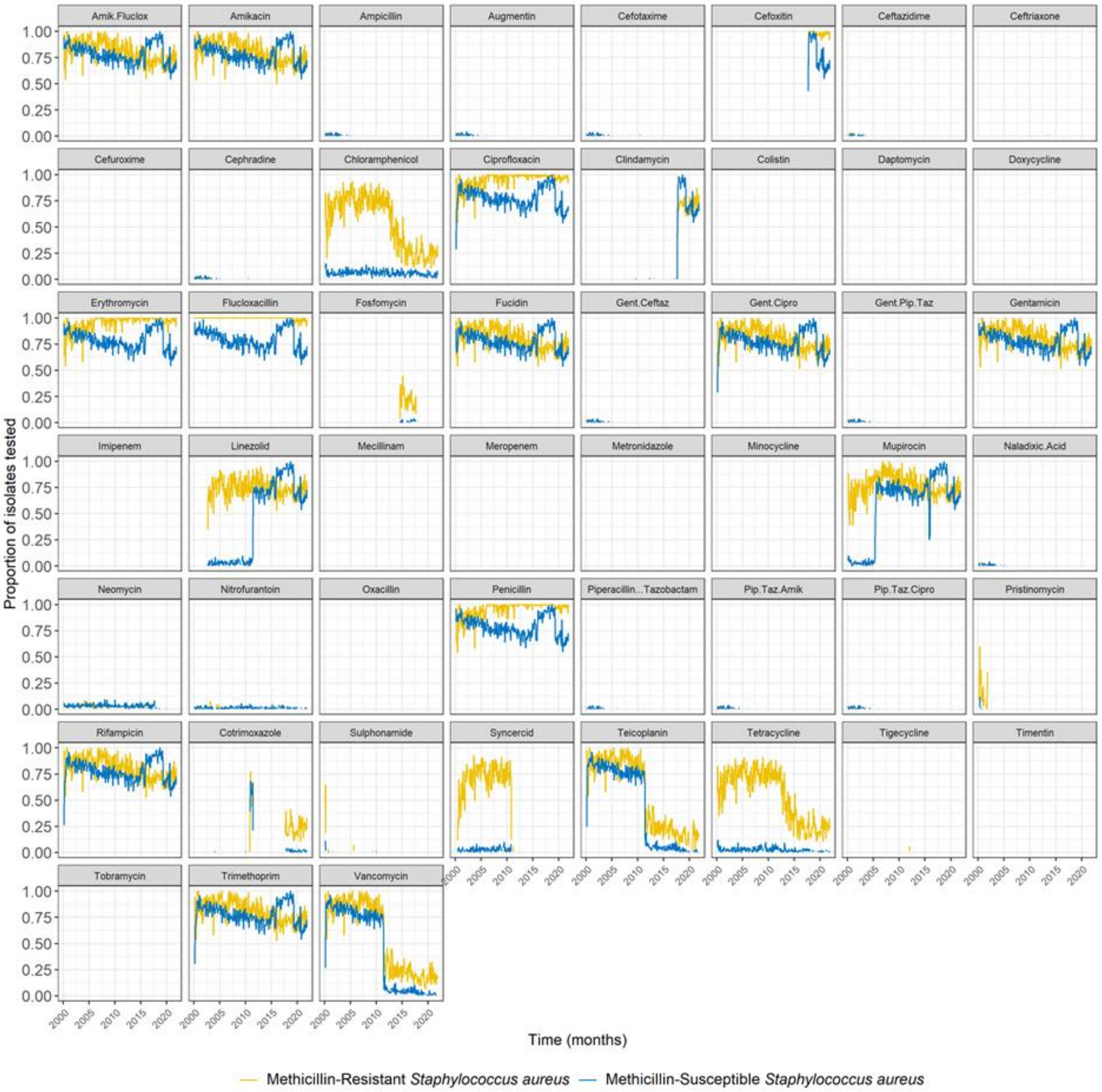


**Supplementary Figure 7: Change in proportion of *S. aureus* isolates tested for different antibiotic susceptibilities over time.** Amik.Fluclox: joint amikacin and flucloxacillin resistance, Gent.Ceftaz: joint gentamicin and ceftazidime resistance, Gent.Cipro: joint gentamicin and ciprofloxacin resistance, Gent.Pip.Taz: joint gentamicin and piperacillin-tazobactam resistance, Pip.Taz.Amik: joint piperacillin-tazobactam and amikacin resistance, Pip.Taz.Cipro: joint piperacillin-tazobactam and ciprofloxacin resistance. Missing lines indicate that no susceptibility testing for the corresponding antibiotic was conducted at that time.

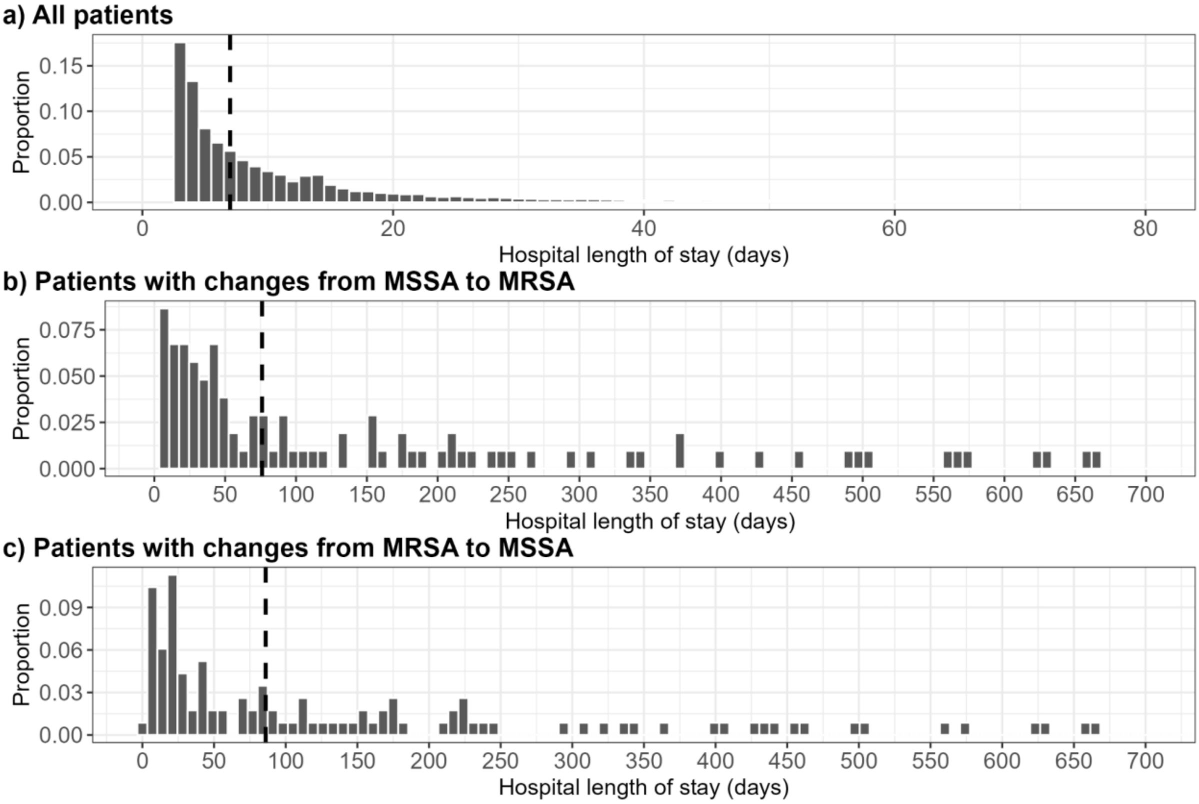


**Supplementary Figure 8: Distribution of hospital length of stay (LoS) for patients in our dataset.** Some patients may have multiple, separate hospital stays. **a) LoS for all patients.** The dashed line shows the median LoS (7 days). Bin size is 1 day. LoS smaller than 2 days were not included. We excluded 1,342 LoS (3.4%) greater than 80 days from the figure for ease of visualisation. **b) LoS for patients with a change from MSSA to MRSA.** The dashed line shows the median LoS (67 days). Bin size is 7 days. We excluded 4 LoS (3.8%) greater than 700 days from the figure for ease of visualisation. **c) LoS for patients with a change from MRSA to MSSA.** The dashed line shows the median LoS (86 days). Bin size is 7 days. We excluded 5 LoS (4.3%) greater than 700 days from the figure for ease of visualisation.

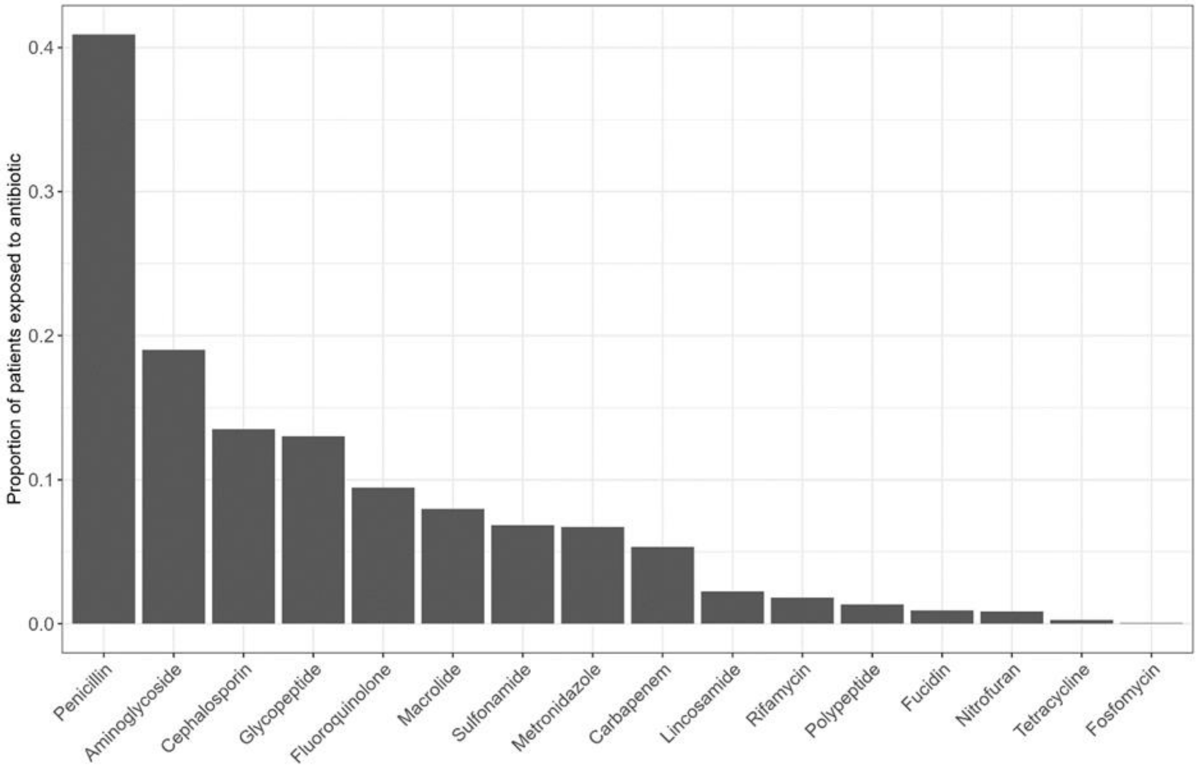


**Supplementary Figure 9: Proportion of all patients with a positive *S. aureus* swab exposed to antibiotics of different classes.** Patients may have been exposed to more than one antibiotic.
